# Mathematical Models for Assessing Vaccination Scenarios in Several Provinces in Indonesia

**DOI:** 10.1101/2020.12.21.20248241

**Authors:** N. Nuraini, K. Khairudin, P. Hadisoemarto, H. Susanto, A. Hasan, N. Sumarti

## Abstract

To mitigate more casualties from the COVID-19 outbreak, this study assessed optimal vaccination scenarios, considering some existing healthcare conditions and some assumptions, by developing *SIQRD* (*Susceptible-Infected-Quarantine-Recovery-Death*) models for Jakarta, West Java, and Banten, in Indonesia. The models included an age-structured dynamic transmission model that naturally could give different treatments among age groups of population. The simulation results show that the timing and period’s length of the vaccination should be well planned and prioritizing particular age groups will give significant impact on the total number of casualties.

## 1 Introduction

In March 2020, the World Health Organization has declared the COVID-19 outbreak as a global pandemic. As of October 2020, it was reported that at least 42 million people had been infected worldwide, with death toll more than one million [1]. By implementing well-planned and systematic mitigation strategies, some countries have been succeeded in suppressing the number of active cases. By contrast, the number of active cases in Indonesia keeps increasing. The death toll in Indonesia is as high as nineteen thousand people [2], which is the highest among southeast Asian countries [3]. The recent update gives even more frustrating situation due to media news reporting some reinfection of COVID-19 [4].

One way to mitigate the COVID-19 outbreak is to develop its vaccine and produce it massively for all affected countries. A vaccine could prevent a susceptible person from being infected at least for a time period or even for life. Recently, several vaccine candidates are being developed around the globe. The SinoVac, a potential COVID-19 vaccine developed by a China-based biopharmaceutical company, has been tested in Indonesia for a Phase 3 clinical trial [5]. This news had brought fresh hope for dealing with COVID-19 mitigation in Indonesia.

However, the existence of the vaccine in the near future is a double-edged sword. Due to the history of other virus-induces illnesses, issues of vaccine mismatch and its suspected side effect on immunocompromised individuals, this existence of vaccine should be well observed. Instead of suppressing the morbidity and mortality of COVID-19, a lack of consideration on vaccination scenarios could also cause several unwanted results, i.e. COVID-19’s second outbreak, ineffective vaccination, etc. This will challenge policy-makers and researchers to consider the best scenario of vaccination for suppressing the mortality and morbidity of COVID-19.

Several studies discussing models on COVID-19 vaccination have been conducted recently. Bitsouni et al. [6] discussed the vaccine effectiveness using the *SEIAR* (*Susceptible-Exposed-Infected-Asymptomatic-Recovered*) model based on case of Italy. The study focused on the risk of infection spread, the peak prevalence of infection and the time at which the peak prevalence occured. The paper by Zindoga et al. [7] estimated the effect of social distancing implementation and explored vaccine efficacy scenarios based on case in South Africa.

in order to describe the behavior of COVID-19 spread on several provinces of Indonesia, i.e. Jakarta, Banten, and West Java, we propose mathematical models, based on non-age-structured and age-structured of SIQRD (*Susceptible-Infected-Quarantine-Recovery-Death*) models. The effectiveness of vaccination program was put into the model by considering the proportion of those who likely to recover or immune from the illness after getting vaccinated. Simultaneously, there is a chance of reinfection by allowing a portion number of recovered people to be re-infected after a particular time. By using these constructed models and simulation scenarios, we propose an estimation of optimum vaccination schedule considering the vaccination cost, healthcare capacity, and vaccination capacity per day. Based on the simulation, the effect of the timing to begin vaccination on the mortality and morbidity cases was also observed. Eventually, we consider several scenarios for prioritizing some age groups and evaluate the results.

## 2 Problem Description

The major questions we would like to adress in this paper are:

(Q1) What is the mathematical model that best represents COVID-19 spread in provinces Jakarta, Banten and West Java, Indonesia? Does the reinfection of recovered people urge the increase of spread significantly?

(Q2) What is the effect of vaccination on mortality and morbidity cases of all age groups? When is the optimal timing to begin the vaccination?

(Q3) Can we find an optimum vaccination scenario due to the existing government’s capacities, including the healthcare facility, estimated maximum capacity of vaccination per day? How many people should be vaccinated for mitigating the spread?

(Q4) If we consider the age-structure of susceptible people in those provinces, should we prioritize on some age groups over others?

The answers to these questions may help the policymaker, especially in Indonesia, for planning the vaccination program in the near future.

## 3 Proposed Models and Its Analysis

In this section, models of *SIQRD* representing the impacted populations due to COVID-19 spread are constructed based on non-age and age structures. We modified the *SIRD* model by adding the compartment *Q* representing the number of quarantined people. The standard type of model has been commonly used in modeling of the vaccination of other virus-induced illness such as influenza [8, 20]. We needed to construct the relation among variables based on the observation on the real examined problem.

### 3.1 Non-Age-Structured Model

Assuming the people in *Q* compartment did not become infectious because they have been under supervised by medical personnel. According to the flow diagram in Fig.1, the vaccination of the suspected people would transfer them directly into compartment *R*. The vaccination scenarios would be accommodated by determining a particular function of *v*(*t*). However, people in *R* would be transferred back to *S* due to the chance of reinfection with rate *ζ*. This modification allows the recovered and vaccinated people to be reinfected. The vaccine might not give the perfect protection from infection due to a mismatch problem of the type of virus being used for the vaccine. Thus, instead of defining it as the total number of recovered persons, we defined *R* as the total number of immune people due to both recovery and vaccination.

**Figure 1.**
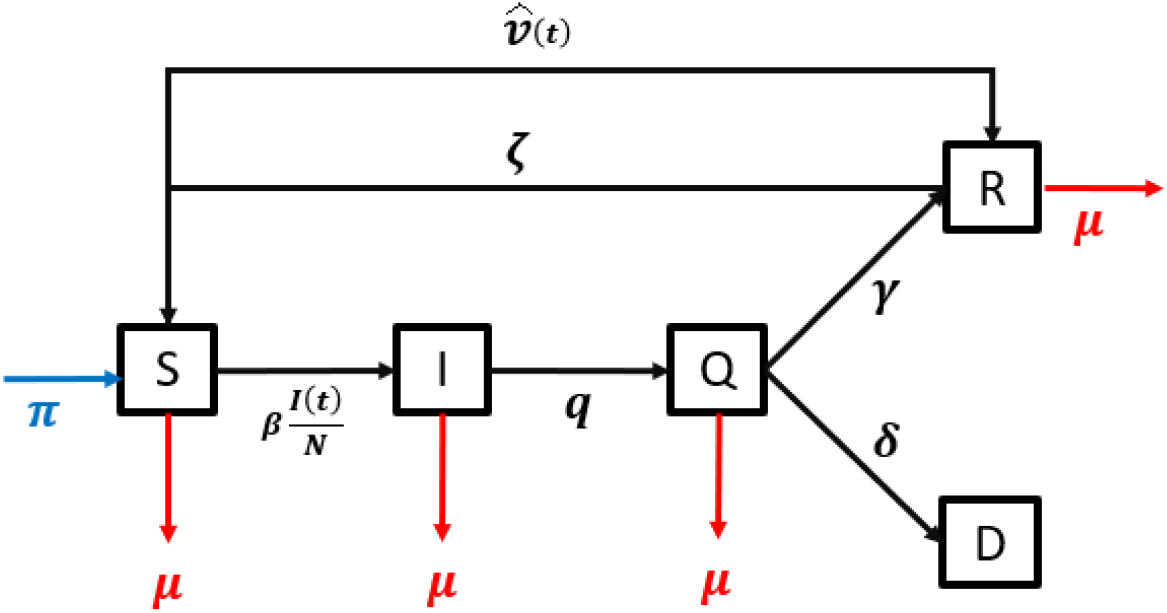
Flow diagram of the SIQRD model. Blue and red arrows represent respectively the natural recruitment and death rate.

The mathematical model is given by

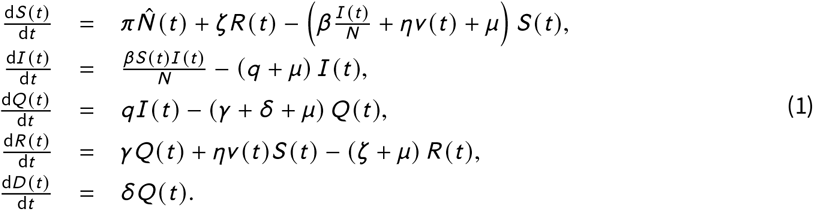

in which system (1) satisfies

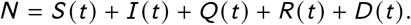

We defined 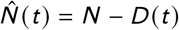 as *π* and assumed to be equivalent to *µ*. The description of Variables and parameters can be found in Table 1.

**Table 1.** Description of Variables and Parameters

| Notations | Description | Unit | Value |
| --- | --- | --- | --- |
| $S(t)$ | Number of susceptible population at time-$t$ | person | - |
| $I(t)$ | Number of infectious population at time-$t$ | person | - |
| $Q(t)$ | Number of quarantined population at time-$t$ | person | - |
| $R(t)$ | Number of immune population at time-$t$ | person | - |
| $D(t)$ | Number of deceased population at time-$t$ | person | - |
| $\beta$ | Transmission rate | 1/day | estimated |
| $\gamma$ | Recovery rate due to COVID-19 | 1/day | estimated |
| $\delta$ | Death rate due to COVID-19 | 1/day | estimated |
| $q$ | Quarantine rate | 1/day | assumed |
| $v(t)$ | Vaccination rate | 1/day | estimated |
| $\eta$ | Vaccine efficacy | - | assumed |
| $\hat{v}(t) = \eta v(t)$ | Potent vaccination rate | 1/day | estimated |
| $\zeta$ | Reinfection rate | 1/day | assumed |
| $\mu$ | Natural recruitment (birth) rate | 1/day | assumed |
| $\pi$ | Natural death rate | 1/day | assumed |

**Table 2.** Parameter estimation of the non-age-structured model for Jakarta

| $\beta$ | $\gamma$ | $\delta$ | $I(t_0)$ | $R_0$ |
| --- | --- | --- | --- | --- |
| 0.4215 | 0.0876 | 0.0028 | 66 | 1.0134 |

### 3.2 Age-Structured Model

The age-structured model of *SIQRD* is similar to the system of equations (1). For each compartment, there are five age groups, i.e. 0-9, 10-19, 20-49, 50-59, and 60 or higher, so all age groups has each compartment assigned to them. For age group *i* ∈ {1, 2, 3, 4, 5},

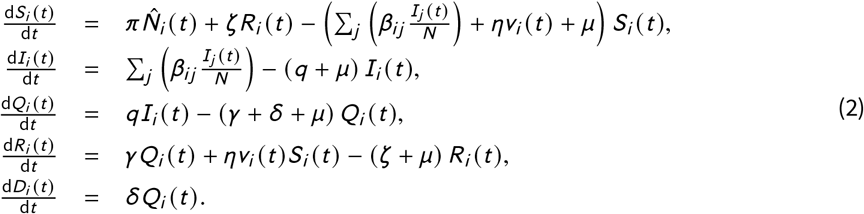

where 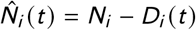, and

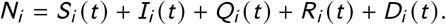

In the first and second equations of system (2), the variable *β*_*i j*_ = *β*_*i*_ *C*_*i j*_ gives cross relationship among age groups, where *β*_*i*_ represents the probability of infection in age group *i*, and *C*_*i j*_ is the matrix giving contacts between any pair of age group *i* and *j*, where *i, j* = 1, 2, …, 5. Here the unit of *C*_*i j*_ and *β*_*i j*_ is 1/day. Function *v*_*i*_ (*t*) gives the potent vaccination rate of group-*i*, which refers to the success of vaccination based on its efficacy for age group-*i*. We assumed the values of *C*_*i j*_ followed the results from the study by Moosong [9], and the values of *β*_*i*_ will be estimated using the provided data of Indonesia.

### 3.3 Vaccination Process in the System

In the constructed models, the vaccination process impacts on a direct transfer of people from compartment *S* to compartment *R* due to the emerging of immune system inside the body of vaccinated people. To develop the vaccination process, we introduced a periodical schedule, where the vaccination was given several times in certain timings so the vaccination rate will be constant between two timings. There will be *k* times of vaccination so there will be *k* periods from the beginning of scenario.

Having determined this definition, *v*(*t*) will be a piecewise function that has constant values for each period of time. The mathematical definition of vaccination rate *v*(*t*) is as follows:

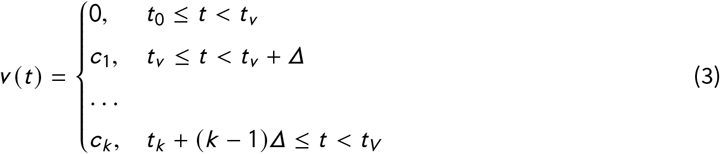

where *c*_*j*_ is a positive constant, *t* _0_ is the initial time of the pandemic, *t*_*v*_ is the time of first vaccination shots, *t*_*V*_ = *t*_*v*_ + *kΔ* is the end of *k* -th vaccination period, and *Δ* is the length of a period. Firstly, the constructed models will be simulated inside the first period before *t*_*v*_ when there is no the vaccination program, or *v*(*t*) = 0 and the result will become the reference used to make the comparison. In this paper, we proposed scenarios of vaccination program by determining the timing *t*_*v*_ + (*j* − 1)*Δ, j* = 1, 2, …, *k* and analyzing the simulation results. The definition of *v*(*t*) was also applied to the age-structured model where the vaccination rate *v*_*i*_ (*t*) refers to the vaccination rate of the age group-*i*. For example, *v*_2_(*t*) is defined the vaccination rate of the 2^*nd*^ age group, which has values *c*_*i j*_ are positive constants, for *i* = 1, 2, …, 5 and *j* = 1, 2, …, *k*

## 4 Datasets

Data of COVID-19 victims of provinces Jakarta, Banten, and West Java was retrieved from https://-19.id/ [2]. It consisted of time-series data of active cases, total recovered cases, and total deaths from late March until late October 2020, which is called as *Dataset 1. Dataset 1* will be used to extract the parameters of the systems (1) and (2). It is an unfortunate that the age-structured COVID-19 data for those provinces was unavailable. In order to estimate parameter *β*_*i*_ in system 2, we made comparison of the population pyramid between each province and a state in USA. Then, it was concluded that data of Connecticut, USA, resembled them enough. We have retrieved COVID-19 data of Connecticut, USA from https://data.ct.gov/ [10]. This website provides time-series data on total infections by age group, which is called as *Dataset 2. Dataset 2* is used to estimate the value of *β*_*i*_ of those provinces. Data on population by age group in provinces Jakarta, Banten, and West Java, and age groups in Connecticut were respectively retrieved from [11, 12] and [13].

## 5 Numerical Results

### 5.1 Parameters Estimation

As in Table 1 for systems (1) and (2), values of several parameters can be estimated using the data, while the rest were based on assumption. The natural recruitment and the natural death rate will be assumed as 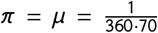, and the life expectancy in Indonesia is about 70 years, based on [14]. The other assumed values of parameters are *q* = 0.4 (the quarantine rate), 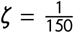 (reinfection rate) according to [15], and *v*(*t*) = 0 as the vaccine is still not available. Values of parameters *β* (transmission rate), *γ* (recovery rate), and *δ* (death rate) will be extracted from the data. The initial number of infected people *I*(*t* _0_) for each province is also estimated. Later, we set *η* = 80% (the vaccine efficacy) when the vaccine is available.

We defined two additional variables, *CR*(*t*) and *V*(*t*), representing the numbers of recovered people and vaccinated people at time-*t*, respectively. The first variable is defined as follows.

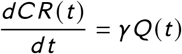

The unknown values and initial conditions of variable *CR*(*t*) are estimated by the Least Square Method (LSM) in Matlab program, so the values of *Q*(*t*), *CR*(*t*) and *D*(*t*) generated from system (1) are close enough to the respective real data of active cases (*DAT A*_*Q*_), total recovered (*DAT A*_*CR*_), and total death (*DAT A*_*D*_). In other words, the unknown values and initial conditions are obtained by solving

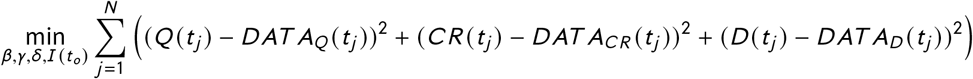

subjecting to *β, γ, δ, I*(*t* _0_) ≥ 0 and *N* denotes the length of data.

Figures 2 and others in the Appendix showed the numerical solutions which performed well in fitting the real data of each province. The second variable was defined using the following differential equation.

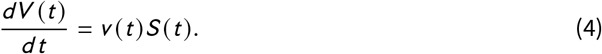

**Figure 2.**
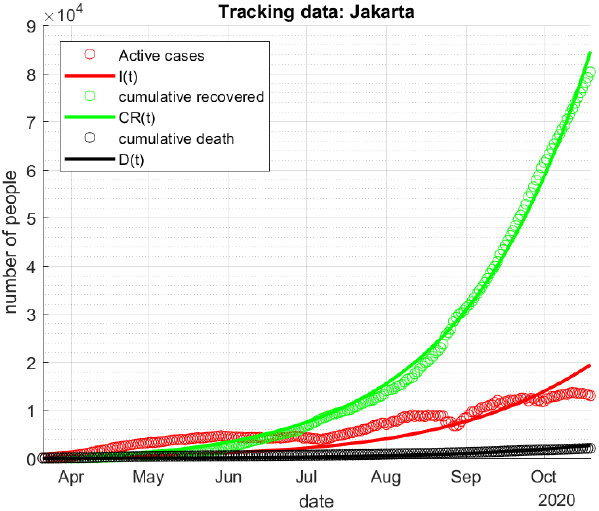
Fitting result of the non-age-structured model for Jakarta

Notice that the number of vaccinated people *V*(*t*) at time *t* was not always fully added up to the number of recovered people *R*(*t*) because of the effect of vaccine efficacy. The values *V*(*t*) are computed as the results of solving its differential equation (4) together with SIQRD model, using the built-in function in MATLAB, i.e. ode45.

To construct the age structured model in system (2) for each province, we applied Connecticut data by using Algorithm 1. Notice that we already have estimated values *β, γ, δ* and *I*(*t* _0_) for the non-age-structured model for each province.

#### Algorithm 1 Computing *β*_*i*_

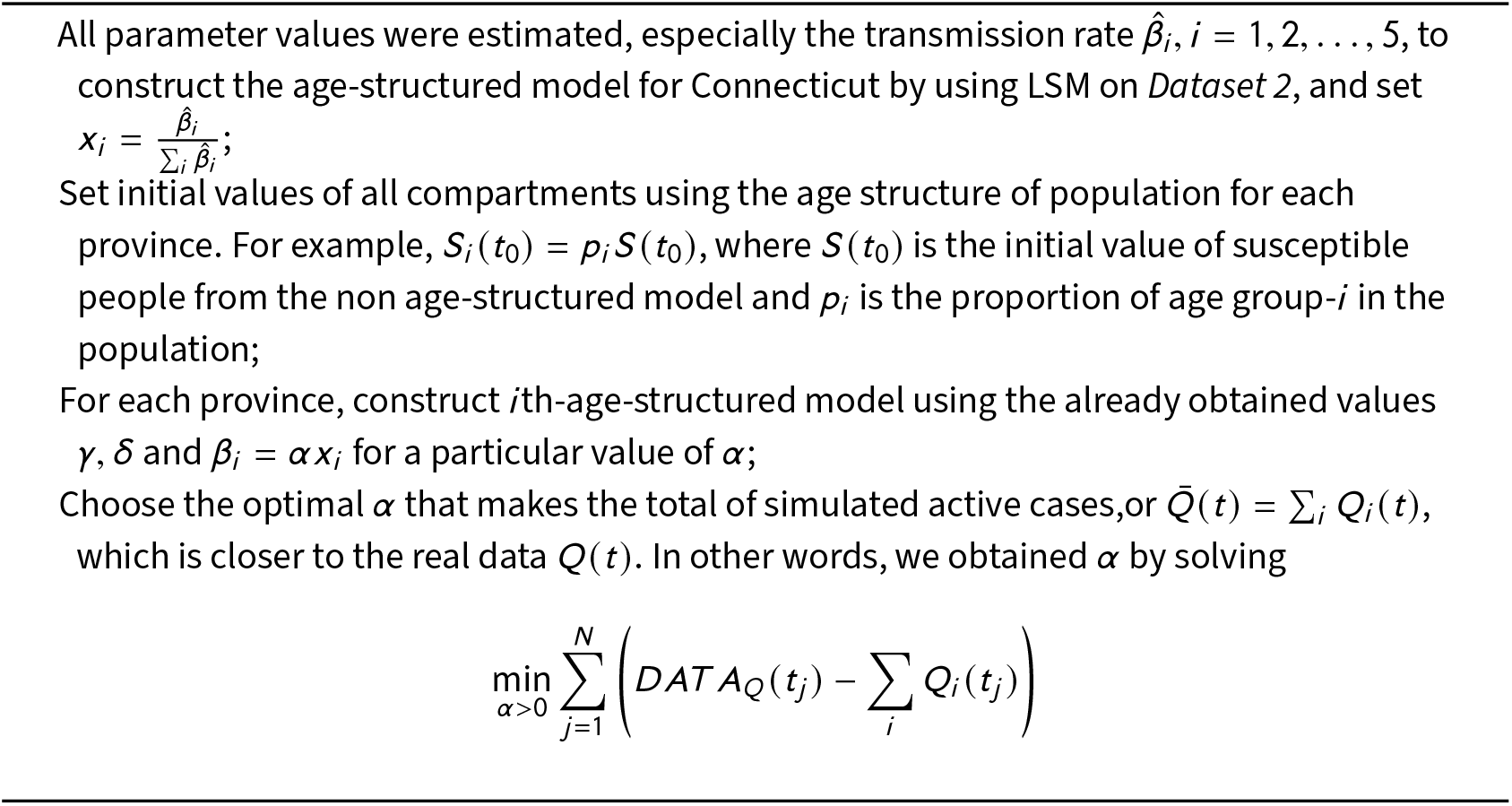

The obtained estimation values of *β*_*i*_ of Jakarta, for *i* ∈ {1, 2, 3, 4, 5} are shown in Table 3. The largest values are owned by age-groups 3,4, and 5 using Algorithm 1. The numbers of active cases in Jakarta are dominated by three older age-groups as seen in Fig.3. The results for other provinces are available in the Appendix.

**Table 3.** Obtained values *β*_*i*_ for the age-structured model of Jakarta

| $\beta_1$ | $\beta_2$ | $\beta_3$ | $\beta_4$ | $\beta_5$ |
| --- | --- | --- | --- | --- |
| 0.0251 | 0.0355 | 0.1122 | 0.1698 | 0.4488 |

**Figure 3.**
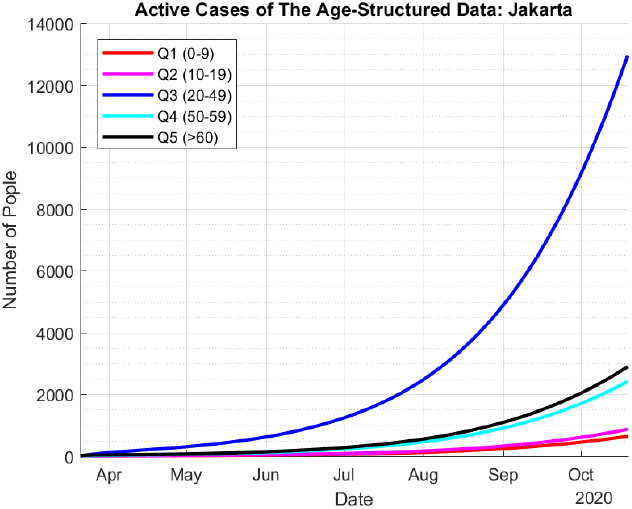
Estimated Active Cases among age groups for Jakarta

### 5.2 Dynamics of Models with and without Vaccination

Having developed the *SIQRD* model of both non-age-structured and age-structured, first we analyzed the dynamics of all obtained models for *v*(*t*) = 0 or without vaccination program, therefore we can project the peak of outbreak that may happen in the near future. Fig.4(a) portrayed the dynamics of *Q*(*t*), *R*(*t*) and *D*(*t*) for non age-structured model of Jakarta. It showed that the peak of active cases will be in February 2021. On the other hand, the number of immune people will largely increase and then decrease as some of them become susceptible again. This condition is possibly causing the second outbreak of the disease later. Notice that for a certain region, the result given by the non-age-structured model was similar to the result given by the age-structured model, e.g. peak occurrence time. However, they are not precisely similar in numbers since we use different objective functions on estimating its parameters. For instance, both Fig.4(a) and (b) are showing that the peak of outbreak will occur on February 2021 in Jakarta. However, they are numerically different since the non-age-structured model estimated that the number of active cases reaches nearly 80,000 cases but the age-structured model estimated 65,000.

**Figure 4.**
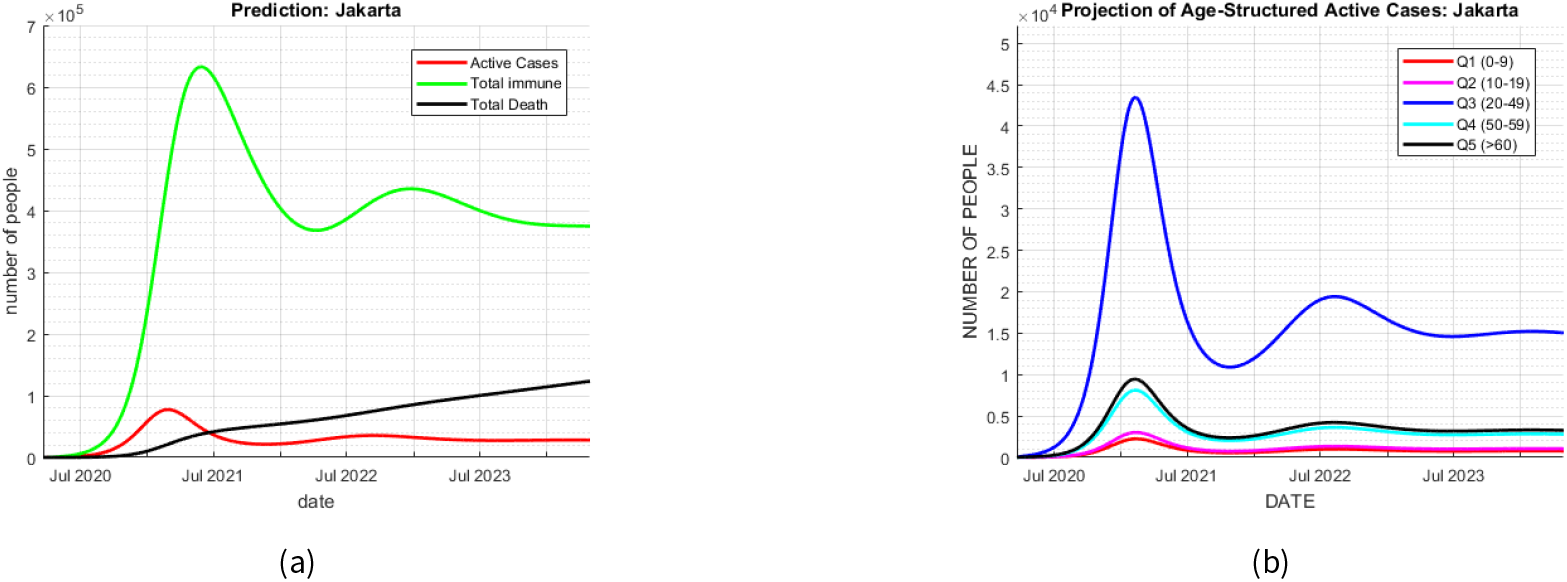
SIQRD simulations without vaccination in Jakarta: (a) non-age-structured; (b) age-structured model

In Fig.4(a), the graph of total number of deaths *D*(*t*) increased because the existence of infectious people. Based on the age-structures model, the dynamics of active cases of Jakarta were classified into 5 age groups in Fig.4(b). Similar to Fig.4(a), the age-structured model also illustrated the second outbreak due to the reinfection for each age-groups. By using the constructed models, this simulation answers (Q1), we can see that the reinfection played a vital role in producing the second outbreak.

The peak of outbreak projections of SIQRD model in Banten and West Java are given by Fig.5. Different from the projection of Jakarta, the dynamics of active cases will increase until August 2021. This is possible because the populations of Banten and West Java are larger than the population of Jakarta. Nevertheless, the general behavior of models is similar where the reinfection factor could cause the second peak of outbreak.

**Figure 5.**
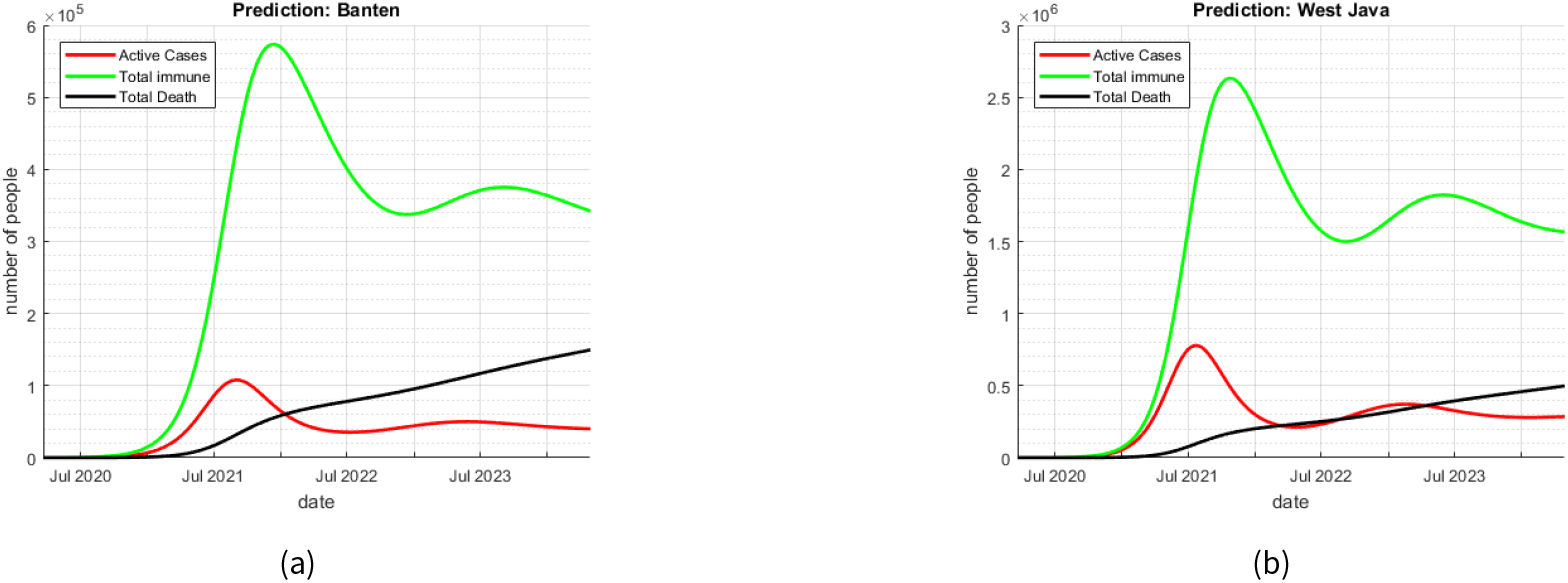
SIQRD simulations without vaccination in: (a) Banten; (b) West Java

Now we discuss whether the existence of vaccination program will generally reduce the number of active cases and total deaths or not. Based on equation (3), we set *k* = 12 and the length of one period was 30 days, so the whole vaccination period was about one year. Set *c*_*j*_ be any values randomly chosen, with 0 ≤ *c*_*j*_ ≤ 10^−3^, which represents the vaccination rate in the *j* th-period, *j* = 1, 2, …, *k*. The vaccination program was set to begin in the third week of October 2020. It is shown Fig.6, the numbers of active cases and total deaths after 1-2 months of vaccination in Jakarta became less than half of the related numbers in the model without vaccination.

**Figure 6.**
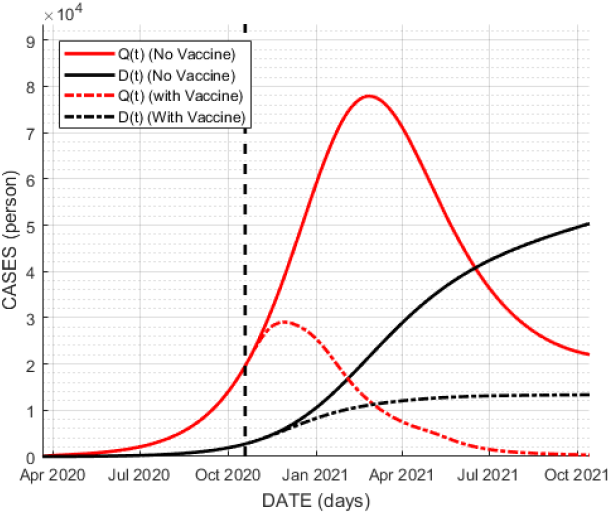
Dynamics of the SIQRD model with vaccination applied in Jakarta

Later in section 6, we will determine the optimal values of *v*_*i*_ (*t*) that met some desired constraints. There will be several scenarios of vaccination being assessed on particular age groups so we could analyse the urgency of vaccination program.

### 5.3 Sensitivity Analysis on Vaccine Efficacy and Quarantine Rate

We wanted to observe the dynamics on the numbers of active cases *Q*(*t*) using simulations with different values of vaccine efficacy *η* and quarantine rates *q*. The first simulation was done by varying the vaccine efficacy and keeping the quarantine rate constant, where the results are in Fig.7(a). The second turn of simulation was using the opposite scheme, where the results are in Fig.7(b). The values for *v*(*t*) were chosen as the same as the ones already used in the previous subsection.

**Figure 7.**
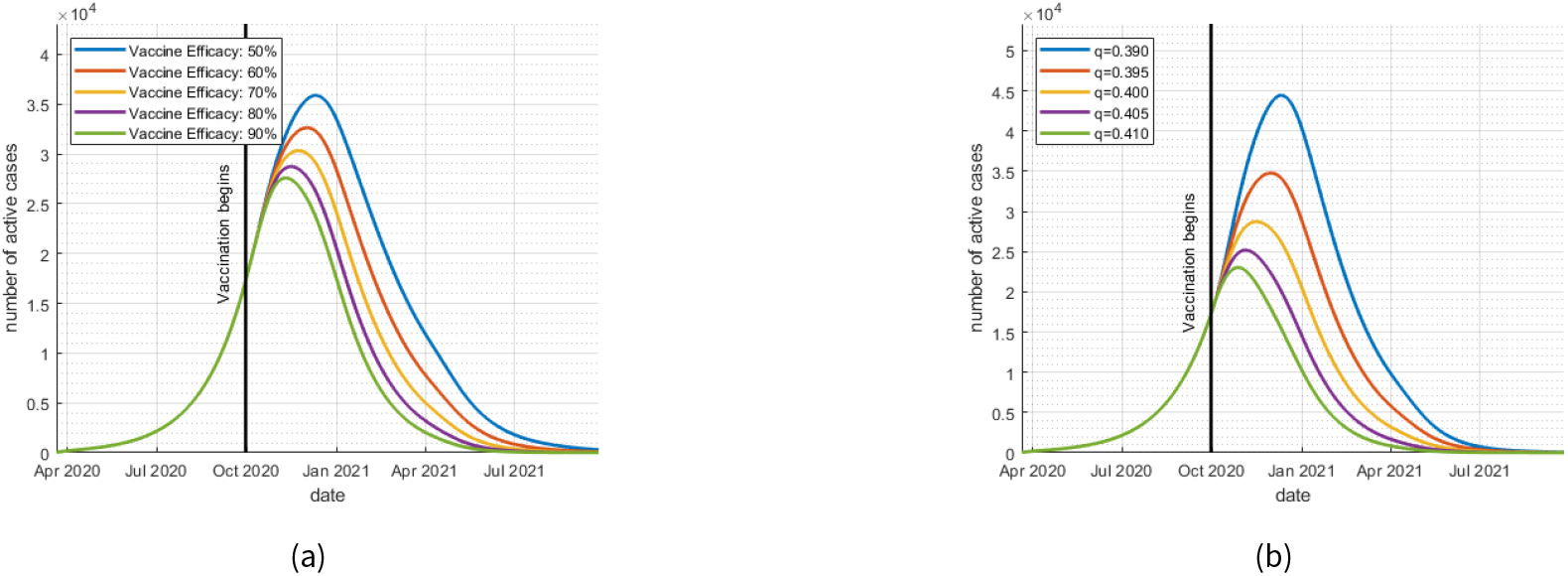
Dynamics of active cases with several values of vaccine efficacy and quarantine rate

Fig.7(a) depicts the change of the numbers of active cases once the vaccine efficacy changed. The greater the vaccine efficacy, the more effective the vaccination is in suppressing the number of active cases. Remember that 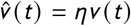. When the vaccine efficacy is low, we require high values of *v*(*t*) to suppress the numbers of mortality and morbidity of COVID-19.

In Fig.7(b), shows that the higher the value of the quarantine rate, the lower the number of active cases is. As a conclusion, we need vaccine with as high efficacy as possible, and also need a quite high quarantine rate to suppress the number of active cases. The total number of deaths will follow the same.

## 6 Several Scenarios of Vaccination

Previously, the simulation of the models was carried out without any constraints in finding the solutions of SIQRD systems (1) and (2). Now we examine several scenarios for vaccination with the main objective of effectively reducing the numbers of active cases and death toll at the minimum cost. It is obvious that the higher vaccination rate needs large number of vaccine provided by the government, and so it requires very high expenses. We established an approximation problem dealing with vaccination function *v*(*t*) with some constraints based on the real situation. There are conditions that should be considered in this optimization process in order to have feasible solution for the real problem, which is as follows:

- The maximum number of active cases does not exceed the maximum capacity of the existing healthcare facility, denoted by *K*_1_ (persons),
- The number of daily vaccine does not exceed the maximum shots of vaccination per day provided by Indonesian government, denoted by *K*_2_ (per day).

The model being observed first was the non age-structured model. Some of results of this optimization problem using this model will be used to execute some scenarios using the age-structured model later. Since comorbidities contribute a high number of deaths, we assumed that comorbidities exist in all age groups. Thus, applying vaccines to all age groups simultaneously can be one of the scenarios worth considering. The scenarios were implemented in finding answer to questions (Q1)-(Q3).

### 6.1 Non Age-Structured Model

In accordance with the required constraints, we defined the optimization problem as follows:

Minimize

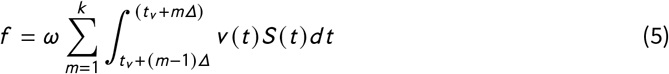

where *t* ∈ [*t*_*v*_, *t*_*V*_] as defined in Section 1. Remember that *Δ* = 30 days is the length of each vaccination period, *k* = 12 is the length of vaccination period, and *ω* represents the expense needed to vaccine a single person.

The objective function *f* has to meet the following constraints:

- max_*t*_ *Q*(*t*) ≤ *K*_1_,
- For *t* ∈ {*t*_*v*_, *t*_*v*_ + 1, *t*_*v*_ + 2, …, *t*_*V*_ − 1},

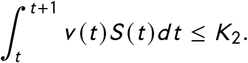
- *v*(*t*) ≥ 0 for *t* ∈ {*t*_*v*_, *t*_*v*_ + 1, *t*_*v*_ + 2, …, *t*_*V*_}

For the maximum number of active cases *K*_1_ and the maximum number of vaccinated persons *K*_2_, we defined two possible conditions; limited and good facilities, and analysed whether there are solutions for this approximation or not.

According to [16], Indonesia government was able to provide about 31.000 COVID-19 testing devices per day nationwide. However these testings kits were not evenly distributed to all provinces in Indonesia. Respectively, Jakarta, Banten and West Java got approximately 31%, 3.4%, and 10% of the total number of provided testing kits. We assumed the limited facility condition based on this news, where the maximum numbers of vaccines received by Jakarta, Banten and West Java were 10.000, 1.100, and 3.100 vaccines per day, respectively. On the other hand, the government claimed there will be 1 million vaccines per day nationwide [17]. We assumed the good facility condition based on this news, so the maximum availability of vaccines were 310.000 vaccines for Jakarta, 100.000 vaccines for West Java, and 34.000 vaccines for Banten.

The simulation using the limited and good conditions of facility was examined when the vaccination starts in October 2020. The above optimization problem with constraint was solved using the built-in function in MATLAB, i.e. *fmincon*. Even after solving the optimum process problem with limited facility condition, solution has not been found yet. So we use the good facility condition for simulations from now onward. In showing the results, the output graph *D*(*t*) is always equipped with the one coming out from the model without vaccination so we can see their stark differences.

#### Starting times of vaccination

We set the vaccination program starting in different timeline; October 2020, January 2021 and after the peak of outbreak for each province. The value *K*_1_ was adjusted for each province to be higher than the existing number of active cases at the time the vaccination starts. The values of *K*_1_ and *K*_2_ are given in Table 4.

**Table 4.** Values of *K*_1_ and *K*_2_ for each provinces.

| Times of vaccination | Jakarta |  | Banten |  | West Java |  |
| --- | --- | --- | --- | --- | --- | --- |
| | $K_1$ | $K_2$ | $K_1$ | $K_2$ | $K_1$ | $K_2$ |
| October 2020 | 30,000 | 310,000 | 15,000 | 34,000 | 50,000 | 100,000 |
| January 2021 | 65,000 | 310,000 | 20,000 | 34,000 | 125,000 | 100,000 |
| After the peak of outbreak | 78,500 | 310,000 | 100,000 | 34,000 | 780,000 | 100,000 |

**Table 5.**
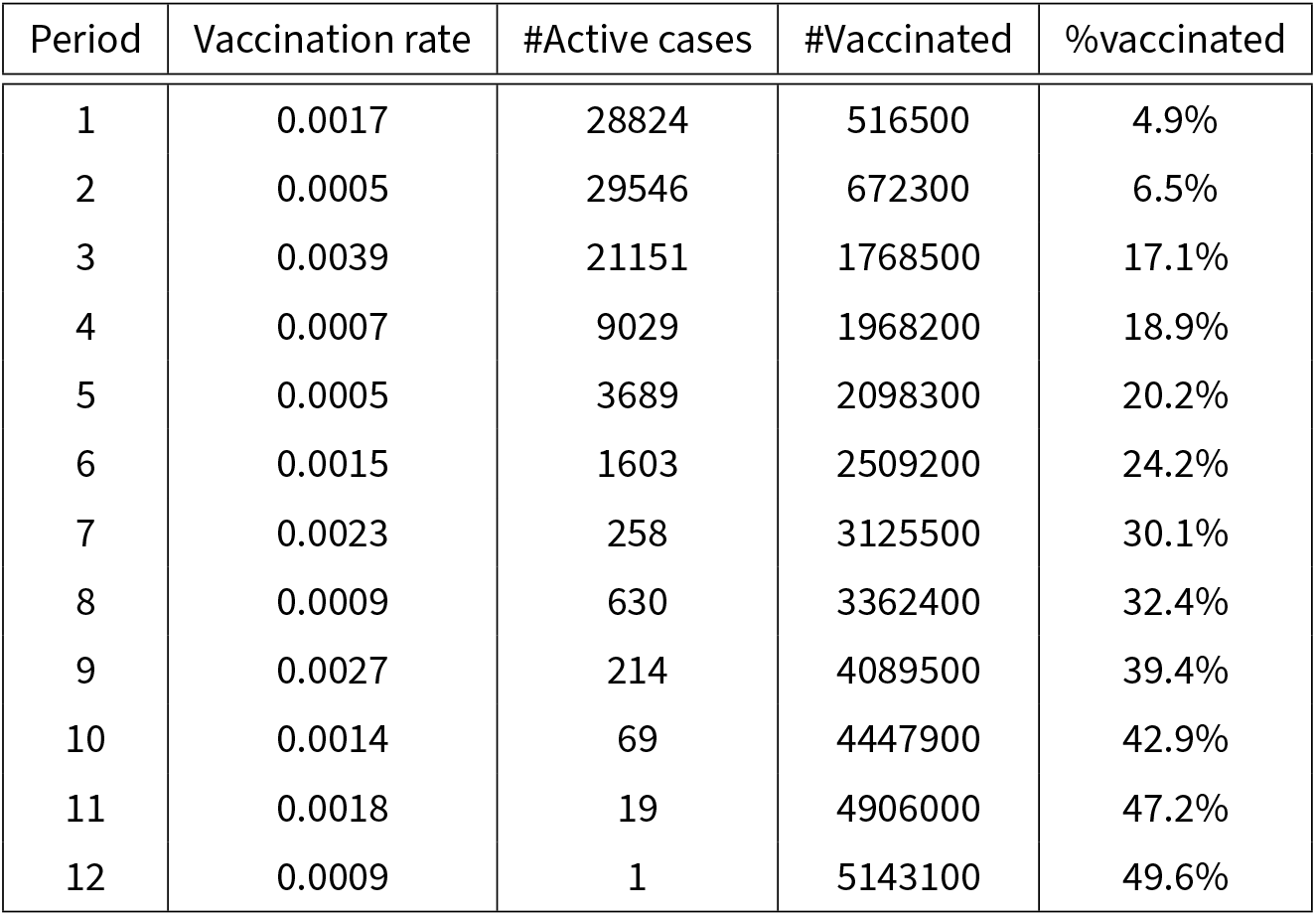
Estimation results of vaccination scheme starting Oct 2020 in Jakarta

| Period | Vaccination rate | #Active cases | #Vaccinated | %vaccinated |
| --- | --- | --- | --- | --- |
| 1 | 0.0017 | 28824 | 516500 | 4.9% |
| 2 | 0.0005 | 29546 | 672300 | 6.5% |
| 3 | 0.0039 | 21151 | 1768500 | 17.1% |
| 4 | 0.0007 | 9029 | 1968200 | 18.9% |
| 5 | 0.0005 | 3689 | 2098300 | 20.2% |
| 6 | 0.0015 | 1603 | 2509200 | 24.2% |
| 7 | 0.0023 | 258 | 3125500 | 30.1% |
| 8 | 0.0009 | 630 | 3362400 | 32.4% |
| 9 | 0.0027 | 214 | 4089500 | 39.4% |
| 10 | 0.0014 | 69 | 4447900 | 42.9% |
| 11 | 0.0018 | 19 | 4906000 | 47.2% |
| 12 | 0.0009 | 1 | 5143100 | 49.6% |

There are many possibilities of obtained values *v*(*t*) coming as the solutions of the optimization problem. Fig.8(a) and (b) are an example of the solution *v*(*t*) that have minimum value of problem (5) for Jakarta. Fig.8(a) shows the graph of *Q*(*t*) that is successfully maintained under the constraint *K*_1_ = 30, 000.

**Figure 8.**
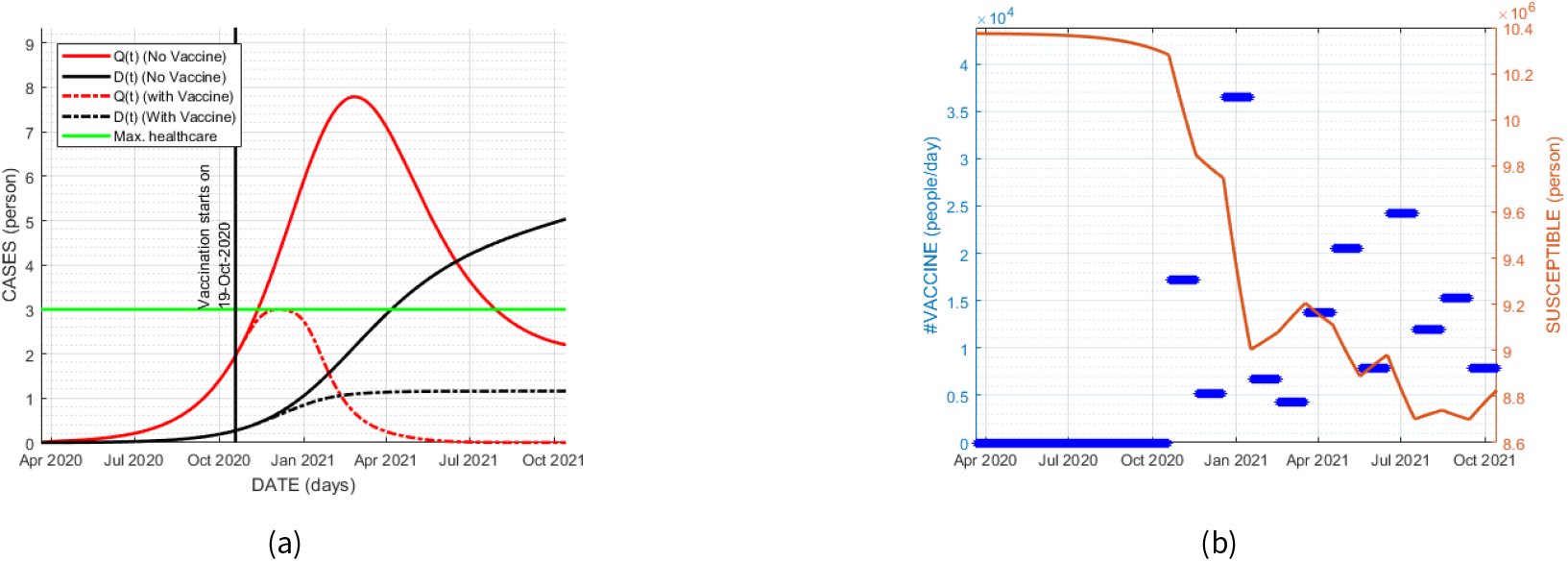
(a) Dynamics of *Q*(*t*) and *D*(*t*) of starting time October 2020 in Jakarta; (b) Number of vaccine per day together with the dynamic of *S*(*t*).

Fig.8(b) shows the number of vaccination per day in Jakarta, represented by the blue thick lines. The number of susceptible people plotted in orange line was drastically decreasing for only after three periods of vaccination. In Table 15, we can see that none of the constraints is violated. The total number of vaccinated people is about 5.14 million, which is only 49.6% of Jakarta’s population. This scenario looks promising where the remaining active cases number is only a single person at the final time *t*_*V*_, or a year later. Unfortunately, this scenario is impossible to apply since in fact the vaccine is not ready yet. Some news media predicted the readiness of the vaccine is not earlier than January 2021.

Fig.9(a) and (b) showed the number of active cases for Banten and West Java. They concluded that the number of people needed to be vaccinated was about 2.9 million people or 23.3 % of population in Banten, and 12.5 million people or 26 % of population in West Java. Tables representing the estimation results of vaccination scheme for Banten and West Java are given in Appendices.

**Figure 9.**
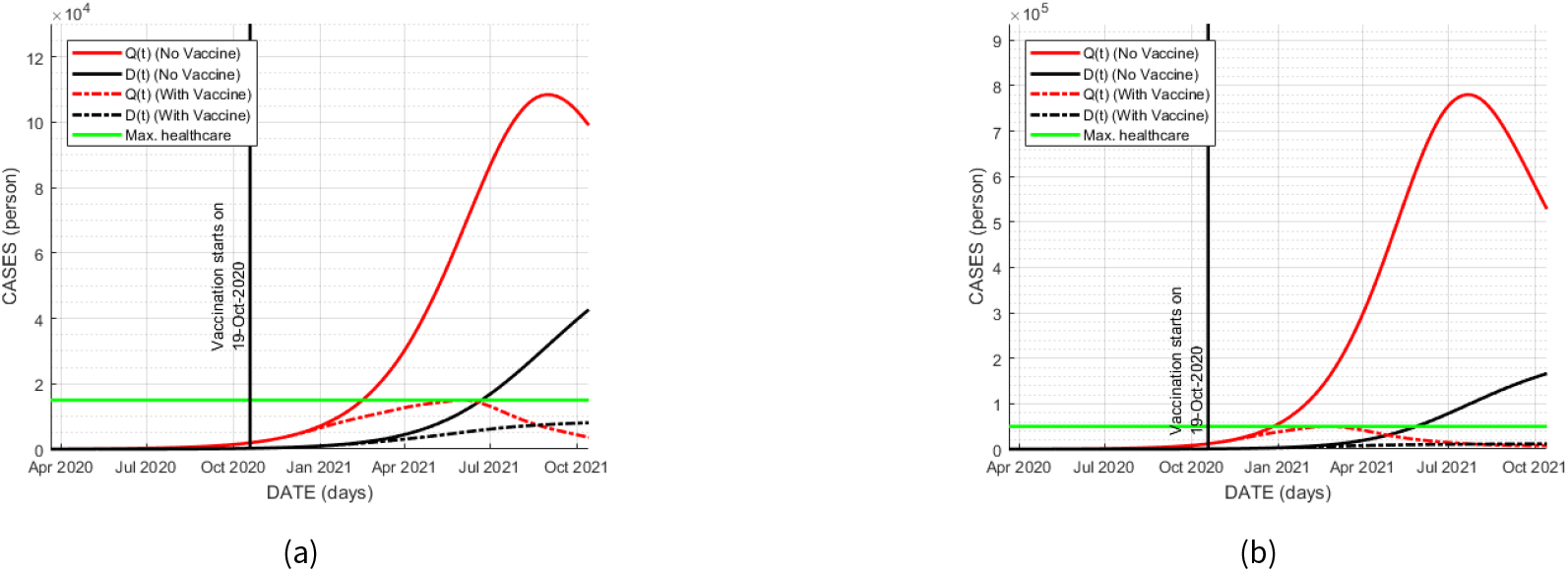
(a) Dynamics of *Q*(*t*) and *D*(*t*) with vaccine on October 2020 in Banten assuming that *K*_2_ = 34.000; (b) Dynamics of *Q*(*t*) and *D*(*t*) with vaccine on October 2020 in West Java assuming that *K*_2_ = 100.000.

If the starting time is in January 2020, Fig.10(a)-(b) and Table 6 give the results for Jakarta. It shows the number of susceptible people rapidly decreased for only after second period of vaccination. The fluctuated number of vaccination per day scheduled for each period, plotted in blue thick line, is one of solutions found from the approximation process. It was interesting to find out whether the rate of vaccination in constant value will be effective or not, because it will be simpler to operate in the real condition. We have this kind of scenario later in this paper.

**Figure 10.**
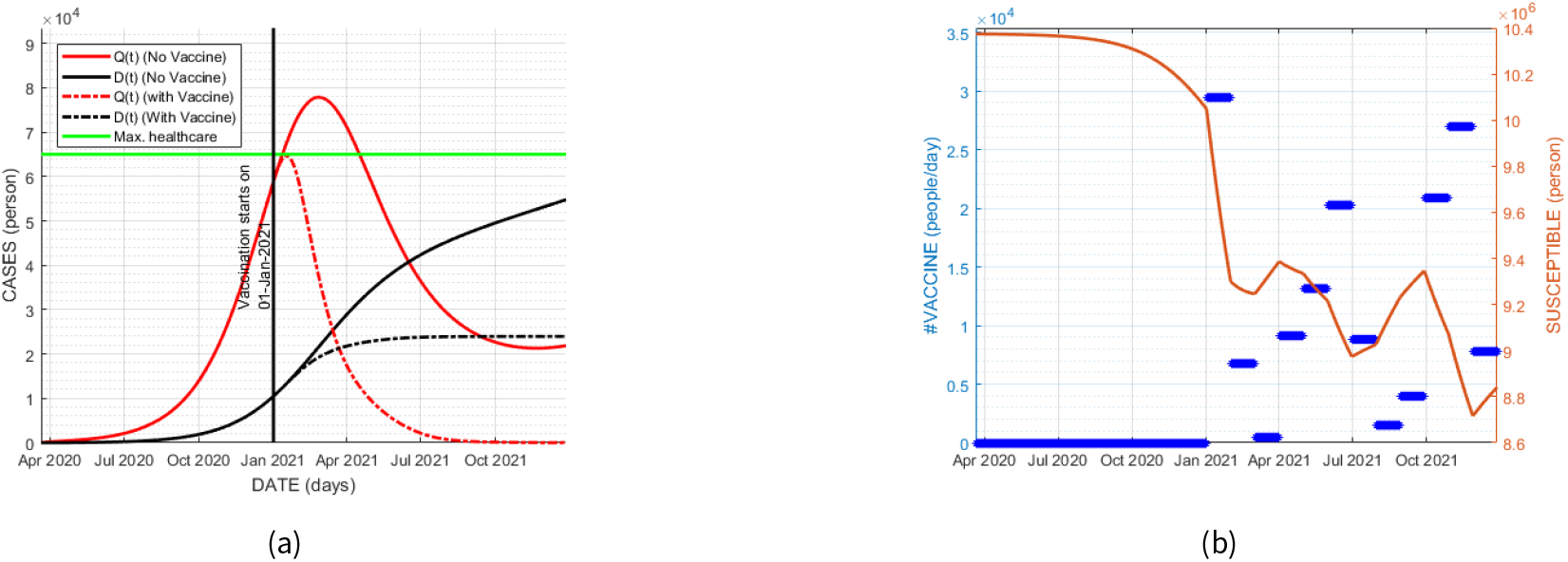
(a) Dynamics of *Q*(*t*) and *D*(*t*) with vaccine on January 2020 in Jakarta; (b) Number of vaccine per day together with the dynamic of *S*(*t*)

**Table 6.** Estimation results of vaccination scheme starting Jan 2020 in Jakarta

| Period | Vaccination rate | #Active cases | #Vaccinated | %Vaccinated |
| --- | --- | --- | --- | --- |
| 1 | 0.0031 | 59515 | 884500 | 8.5% |
| 2 | 0.0006 | 34029 | 1088200 | 10.5% |
| 3 | 0.0001 | 17463 | 1103400 | 10.6% |
| 4 | 0.0010 | 9578 | 1378500 | 13.2% |
| 5 | 0.0014 | 5009 | 1773500 | 17.1% |
| 6 | 0.0022 | 2242 | 2380700 | 22.9% |
| 7 | 0.0010 | 840 | 2646400 | 25.5% |
| 8 | 0.0002 | 326 | 2691200 | 25.9% |
| 9 | 0.0004 | 152 | 2809400 | 27.1% |
| 10 | 0.0023 | 74 | 3437400 | 33.2% |
| 11 | 0.0030 | 28 | 4245100 | 40.9% |
| 12 | 0.0009 | 1 | 4475400 | 43.2% |

From Table 6, the number of vaccinated people was 4.48 million or 43.2% of the population in Jakarta. The vaccination was successful because the remaining number of active cases is only one. The simulation of the vaccination in Banten and West Java are given in Fig.11, where the number of active cases is limited to the value of *K*_1_. The number of vaccinated people was 2.1 million or 17% of the population in Banten, and 10.1 million or 22.7% of the population in West Java.

**Figure 11.**
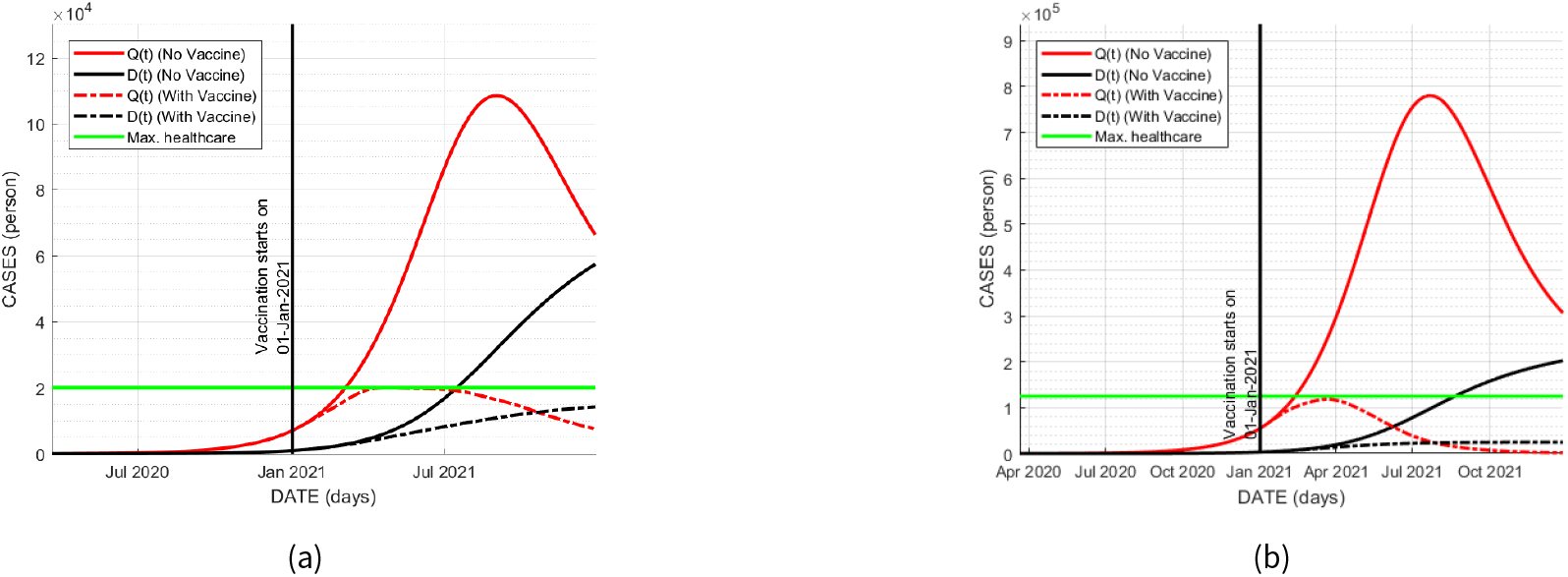
(a) Dynamics of *Q*(*t*) and *D*(*t*) with vaccine on January 2020 in Banten; (b) Dynamics of *Q*(*t*) and *D*(*t*) with vaccine on January 2020 in West Java

Now the starting time of vaccination is after the number of active cases reaches its peak. So the starting time is different for each province, April 2021 in Jakarta, 1.25 million in Banten, and 8.5 million in West Java. Fig.12(a) shows the steeper decrease of *Q*(*t*) than of the decrease without vaccination in Jakarta. The number of death *D*(*t*) seems to stabilize to about 4,000 due to the peak of outbreak happened before the vaccination. From Table 7, total number of people to be vaccinated is 3.19 million or 30.4% of population in Jakarta. If we compare among figures 8(a), 10(a) and 12(a) in sequence, total numbers of death are increasing, which means the later the starting time of vaccination, the higher the number of the pandemic casualties.

**Figure 12.**
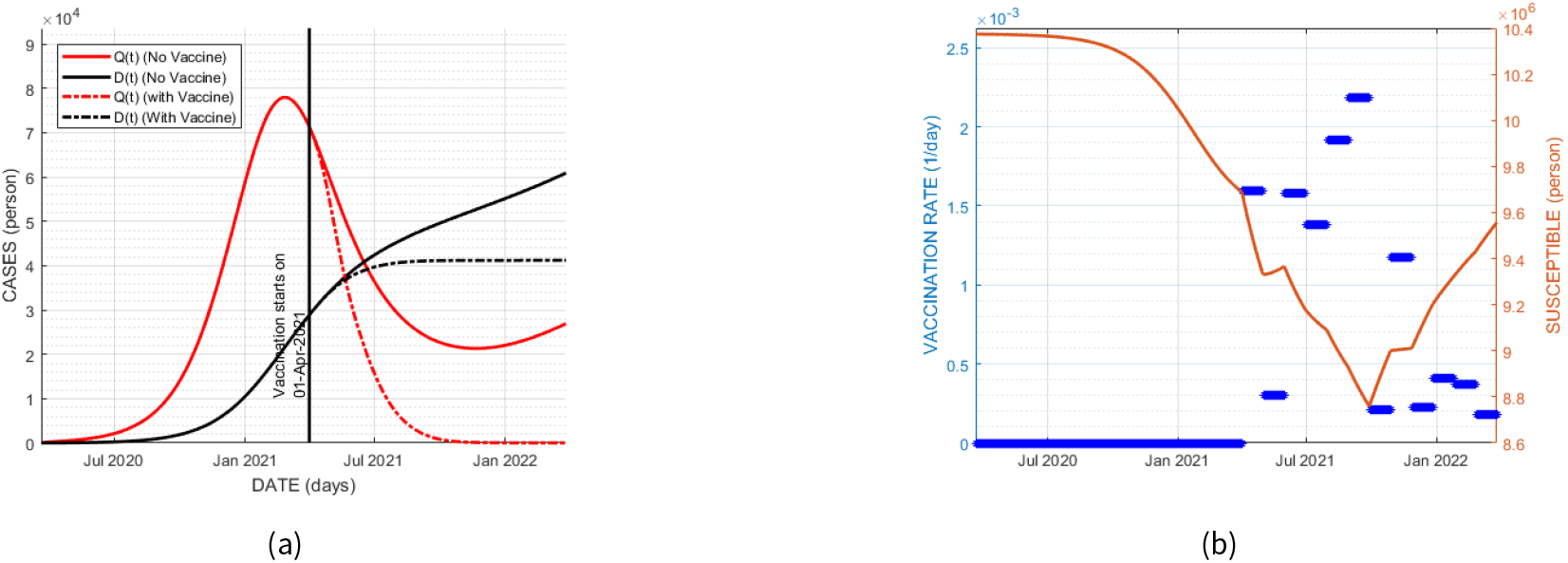
(a) Dynamics of *Q*(*t*) and *D*(*t*) with vaccine on April 2021 in Jakarta; (b) Number of vaccine per day together with the dynamic of *S*(*t*)

**Table 7.** Estimation results of vaccination scheme starting April 2021 Jakarta

| Period | Vaccination rate | #Active cases | #Vaccinated | %Vaccinated |
| --- | --- | --- | --- | --- |
| 1 | 0.0016 | 53327 | 453600 | 4.3% |
| 2 | 0.0003 | 30624 | 537400 | 5.2% |
| 3 | 0.0016 | 16166 | 976900 | 9.4% |
| 4 | 0.0014 | 7285 | 1355800 | 13.1% |
| 5 | 0.0019 | 2863 | 1874500 | 18.1% |
| 6 | 0.0022 | 946 | 2454600 | 23.7% |
| 7 | 0.0002 | 284 | 2510400 | 24.2% |
| 8 | 0.0012 | 96 | 2827000 | 27.2% |
| 9 | 0.0002 | 36 | 2888000 | 27.8% |
| 10 | 0.0004 | 16 | 3001000 | 28.9% |
| 11 | 0.0004 | 9 | 3106000 | 29.9% |
| 12 | 0.0002 | 5 | 3186900 | 30.4% |

#### Frequency of the vaccination

In Fig.10(a), the vaccination for 12 months could make the number of active cases starting to decrease significantly in the first month of vaccination period. It is interesting to see whether one time vaccination in the first month could really work. Most of the time when the first attempt showed good result, vaccination program could be immediately stopped to reduce cost.

Fig.13 depicts a simulation of this one-time vaccination for Jakarta, where *v*(*t*) = 0.0031 for time *t* in January 2020, and *v*(*t*) = 0 for other months. It shows a significant decrease of the active cases at the beginning of vaccination, but it will start to increase from August 2021 onward, so this may cause another outbreak in the future. This prediction is clearly proven when we plot the graphs in longer time-span in Fig.14. On the left, the vaccination consistently given for 12 months from January 2021 can make the non-existence of active cases happen until the beginning of year 2024. On the right, the one-time vaccination potentially impacts on another outbreak in July 2022. The increase of death toll seems to be slowed down about one year, then it will increase to the same number as the obtained number from the non-vaccine model with few-month delay

**Figure 13.**
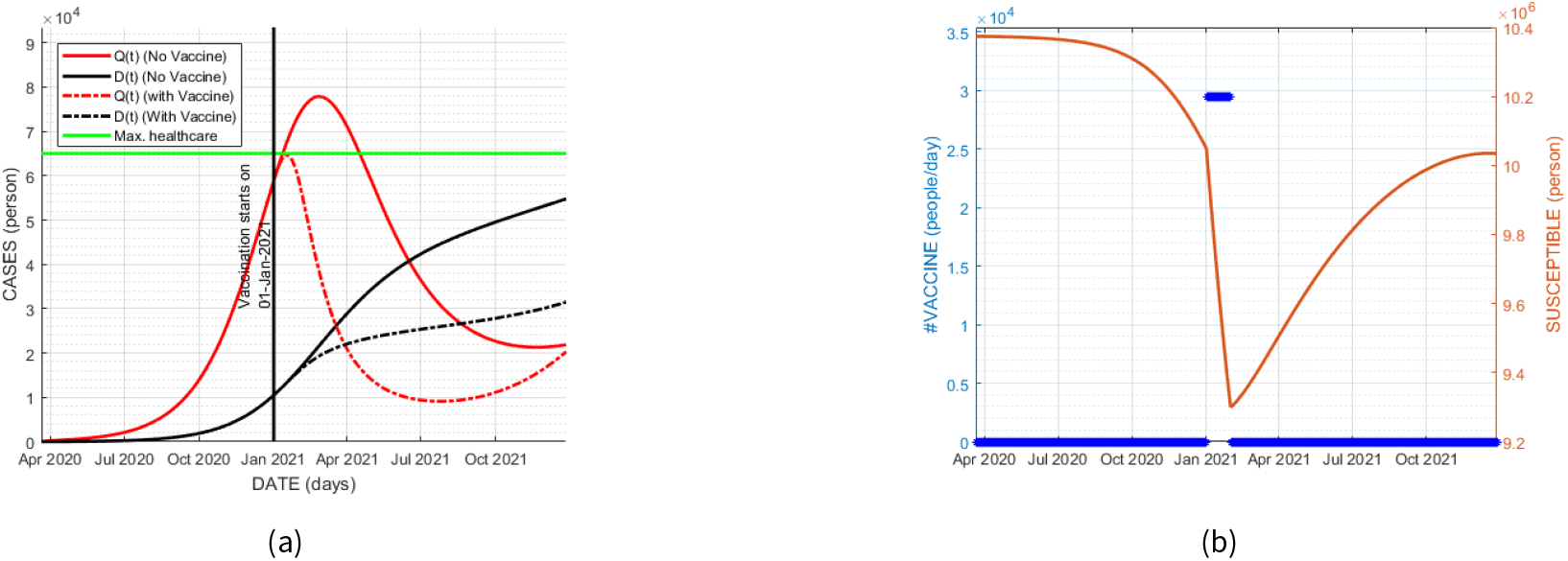
(a) Dynamics of *Q*(*t*) and *D*(*t*) with vaccine only in January 2021; (b) Number of vaccine per day and the dynamic of *S*(*t*)

**Figure 14.**
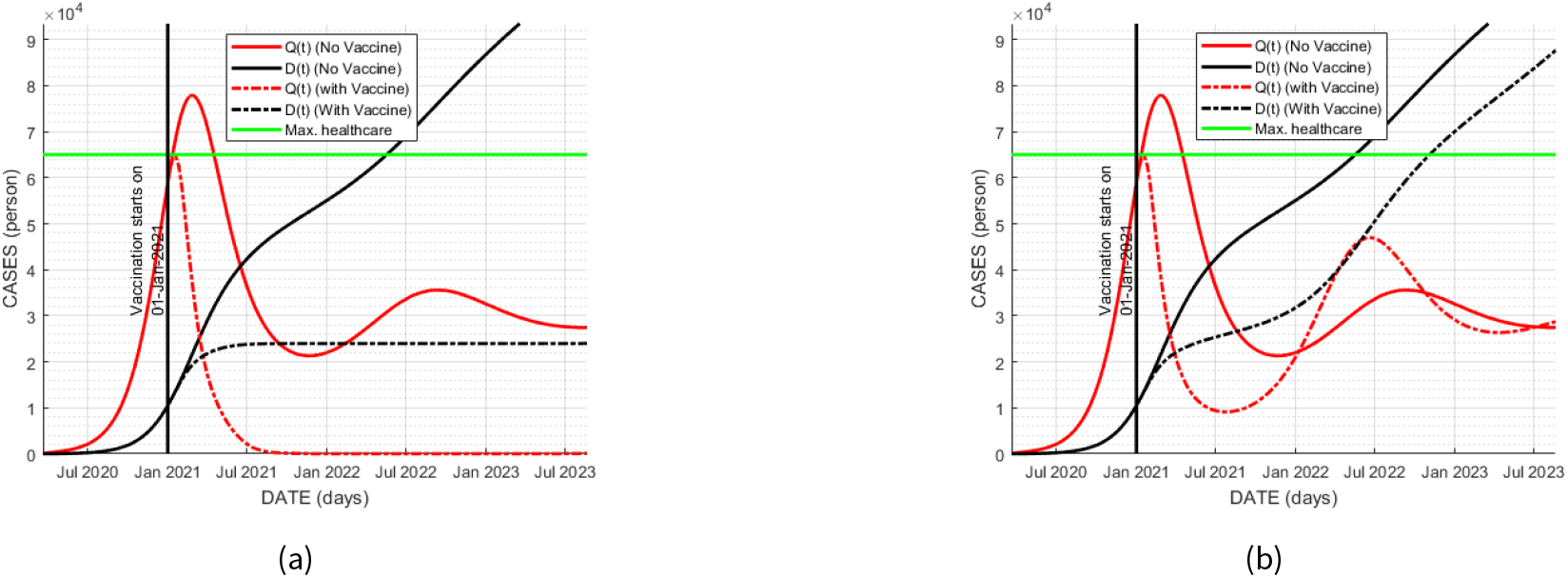
(a) 12-month vaccination; (b) One-month vaccination in Jakarta

#### Constant vaccination rates

The scenarios from the previous subsection assessed the timing and frequency of vaccination. The scheme of vaccination obtained from the approximation process gave fluctuating values of vaccination rates making it hard to be implemented. It means there will be fluctuating needs of the vaccination workers per period. Now we simulate the vaccination with constant rates and analyze the dynamics of the obtained numbers of active cases.

There are 3 scenarios with different values of constant rates starting from January 2021 based on the optimal vaccination rates obtained on Table 6. The constant vaccination rate for the first, second and third scenarios are the average of rates 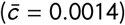, the maximum rate (*c*_*max*_ = 0.0031), and the minimum rate (*c*_*min*_ = 0.0005), respectively. The number of vaccinated people of those scenarios are plotted in Fig.15, where the original optimum values, the average, the maximum, the average and the minimum rates are respectively represented by blue stripes, upper magenta stripes, middle magenta stripes, and lower magenta lines. Due to the decreasing number of susceptible people, the number of vaccinated ones also decreasing, even though the vaccination rates are constant.

**Figure 15.**
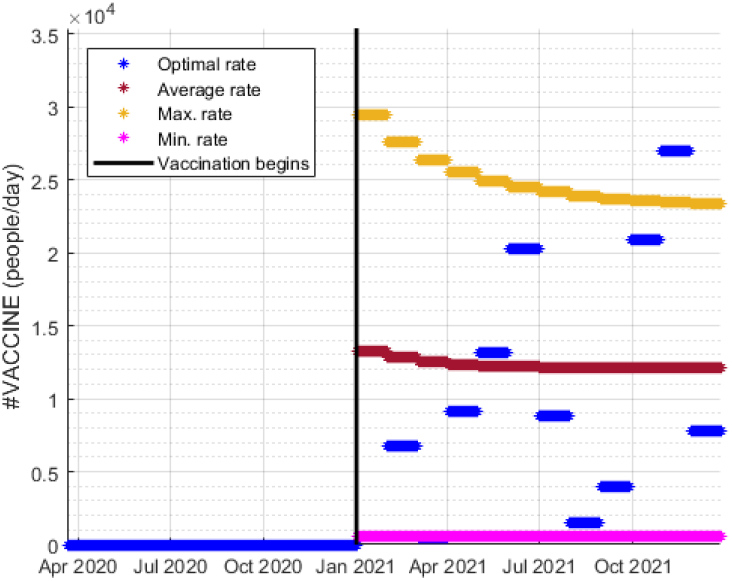
The number of vaccinated people on each period for several schemes: optimum, maximum, average, and minimum of vaccination rate.

Let the optimum fluctuated rates be the benchmark of the uses the healthcare facility that requires an expense calculated from equation (5). The expenses calculated from implementing those scenarios will be compared to this benchmark expense. Fig.16 showed the active cases *Q*(*t*) from all scenarios where the optimum vaccination rate plotted in solid blue line. The average rate scenario gives values *Q*(*t*) exceed the maximum healthcare capacity (the green line) and it requires 99.1% of the benchmark cost. The maximum rate scenario gives plot of *Q*(*t*) that resembles the plot from the optimum rates, but it requires almost twice of the benchmark cost, i.e. 192%. Finally, the minimum rate scenario gives insignificant reduction of *Q*(*t*) compared to the plot of *Q*(*t*) without vaccination. It concludes the optimum fluctuated rates are the best scheme among other constant rate scenarios based on the dynamic of active cases and the required cost.

**Figure 16.**
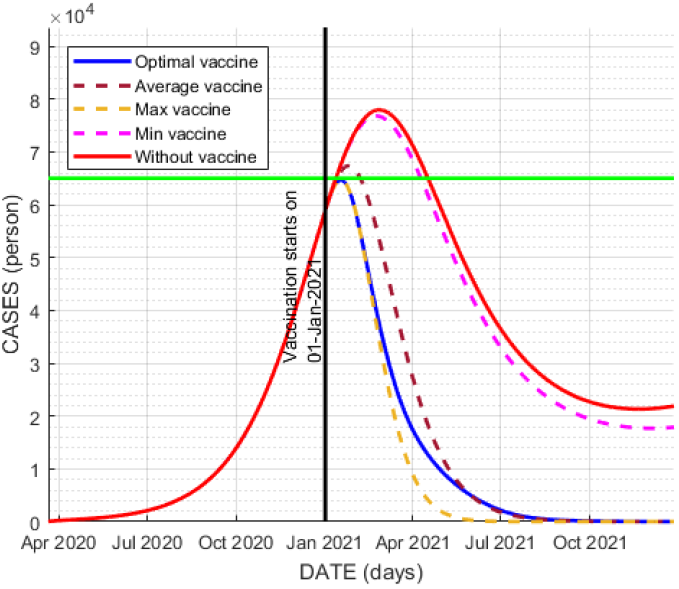
The dynamics of *Q*(*t*) using the constant value of vaccination rate

## Discussion

Considering the results of the simulation of starting time scenarios, the vaccination program ideally should starts in the middle of the pandemic, which is October 2020. The total number of deaths can be suppressed significantly. Unfortunately, this scenario is impossible to apply since the vaccine was not ready yet at that time. If the beginning of vaccination program is delayed as in the scenarios, the required amount of vaccine will decrease, so it seems better to begin vaccination program after the peak of outbreak with the least expense. However, delaying starting time will resulted in increasing total deaths. In fact, the decrease of total number of deaths is not significant if we choose the latest scenario.

Having simulated on the consistency on the frequency of vaccination, it shows in Fig.14(a) that only a single time of vaccination gives insignificant reduction of the number of active cases at the end of the intended vaccination program. Moreover, the final number of infectious people in the simulation, *I*(*t*_*V*_), was about 800 people in Jakarta, which is still too high so it could trigger another peak of outbreak through reinfection. It is concluded that a consistent schedule of vaccination will significantly reduce the number of active cases at the end of vaccination program.

The pattern of the vaccination rate is also interesting to observe. Having seen the results from the first two scenarios, where the vaccine was applied before the number of active cases reaches its peak, the vaccination rate obtained from the optimization procedure was high at the beginning of vaccination schedule. It seems the healthcare facility is not yet at maximum capacity *K*_1_, so much effort can be used to reduce the number of active cases by maximizing the vaccination shots per day. On the other hand, if the vaccine is applied after the peak of outbreak, the vaccination rate will be low at the beginning and then high at the end of the vaccination. In this scenario, the healthcare facility would have been already at the maximum capacity, so the main focus is preventing another peak of outbreak to come.

### 6.2 Age-Structured Model

Now four scenarios of prioritizing particular age groups are implemented on the age-structured model, where the priority is respectively given to groups of the active and older people, groups of 20 years old and younger, group of active people only, or alternating target’s group in each period. The starting time is January 2020 and we use the scheme of vaccination rates shown in Fig.10.

#### Groups of active and older adults (above 50)

During the first six months, the vaccine shots were given to workers and high-risk people, which are 20 and older, then they were given to younger groups in the remaining months. As seen in Fig.17(a), the darker color of the cells for certain periods means higher rate of vaccination. Fig.17(b) illustrates the simulation result of the total numbers *Q*(*t*) and *D*(*t*) of all age-groups using this scenario in Jakarta. The simulation results from Banten and West Java are given in Fig.18. The vaccination performs well in the reduction of the numbers of active cases and death toll.

**Figure 17.**
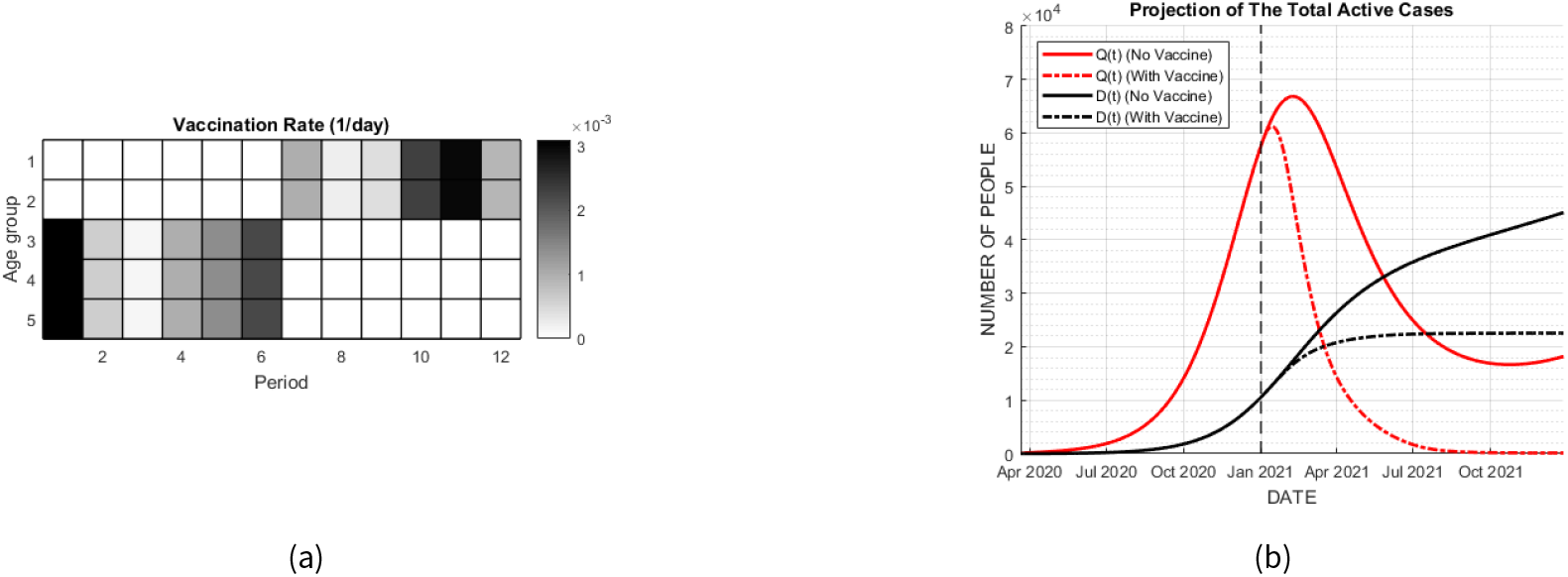
Vaccination scenario by prioritizing active and older adults (above 50) (a) Vaccination schedule; (b) Dynamics of *Q*(*t*) and *D*(*t*) in Jakarta

**Figure 18.**
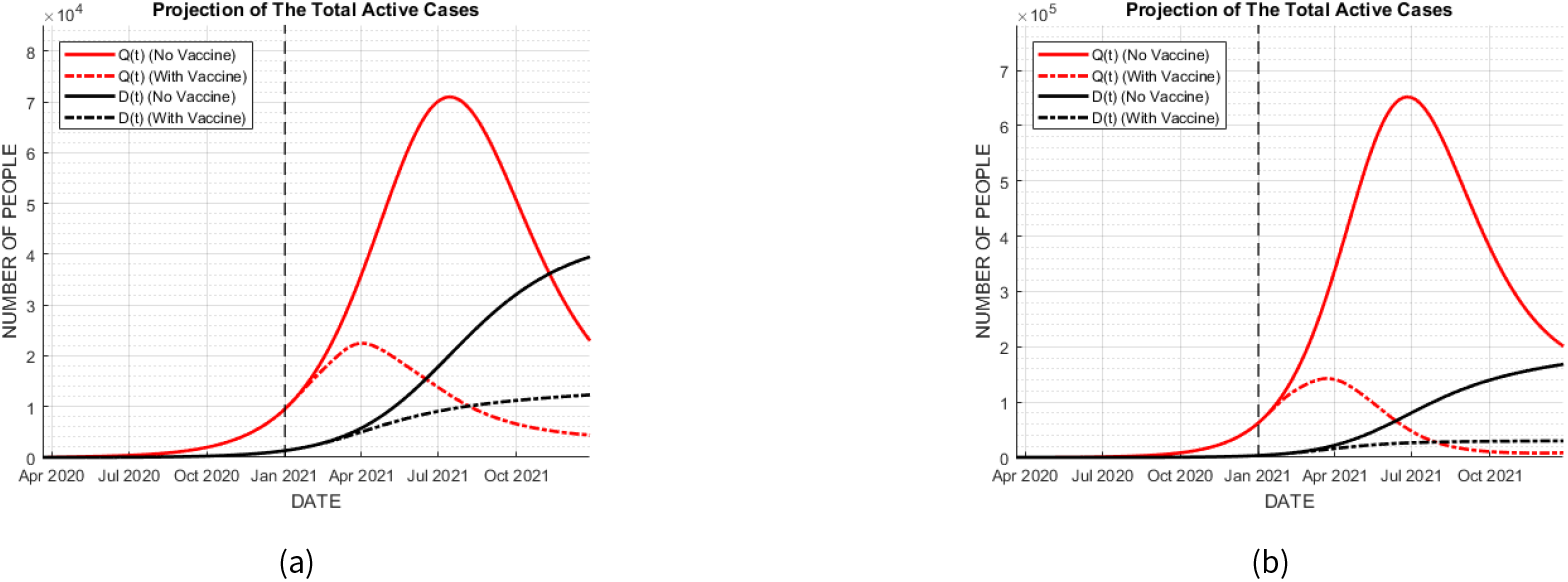
Vaccination scenario by prioritizing active and older adults (above 50) in (a) Banten; (b) West Java.

#### Groups of 20 years old and younger persons

In this scenario, the first interval of six months is the vaccination time only for the younger people with ages 0-19, and the remaining time is for the active and older people’s groups, as scheduled in Fig.19(a). The dynamics of total numbers *Q*(*t*) and *D*(*t*) of all age-groups using this scenario in Jakarta, Banten and West Java are described in Fig.19(b) and Fig.20, respectively. It seems this scenario gives insignificant decrease on the total numbers of active cases and death toll.

**Figure 19.**
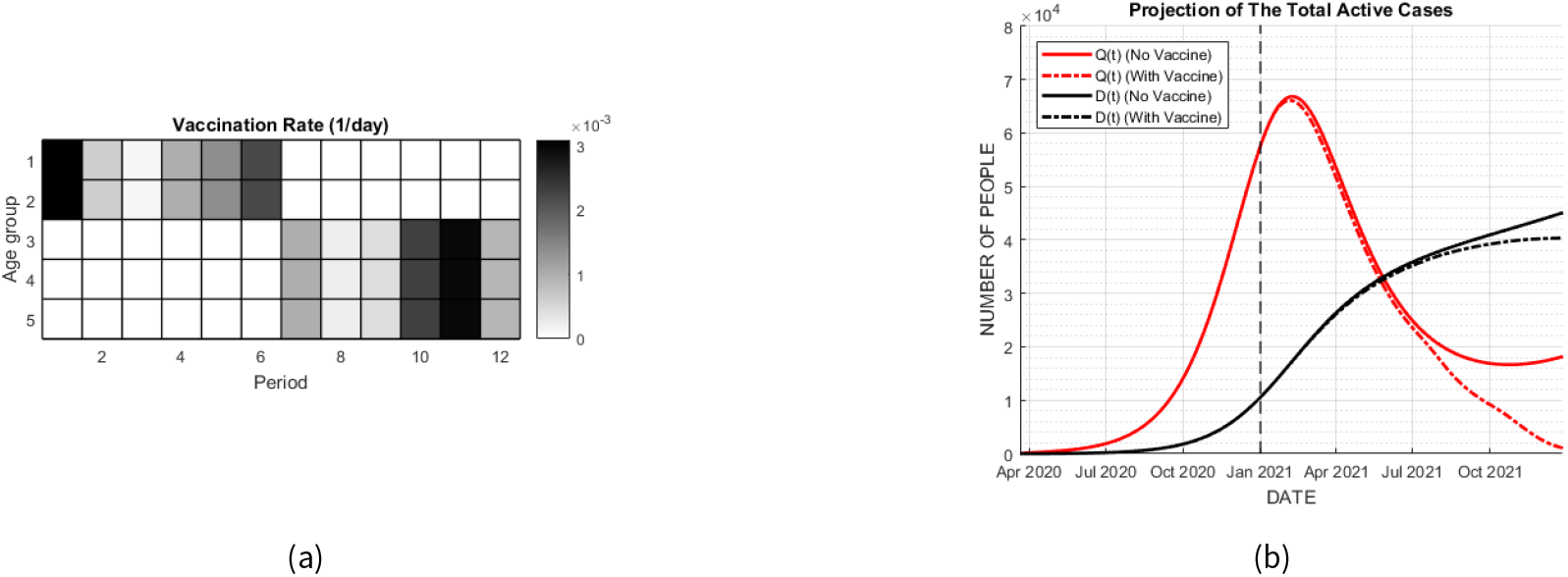
Vaccination scenario by prioritizing 20 years old and younger (a) Vaccination schedule; (b) Dynamics of *Q*(*t*) and *D*(*t*) in Jakarta

**Figure 20.**
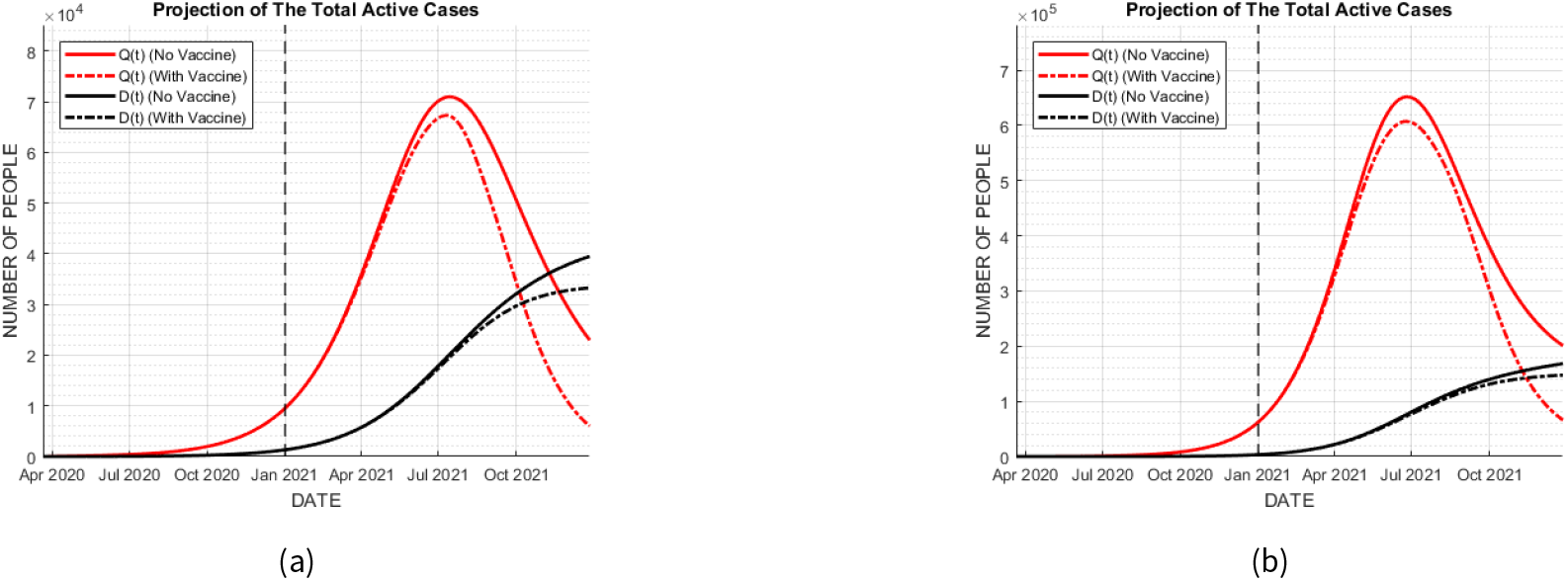
Vaccination scenario by prioritizing 20 years old and younger in (a) Banten; (b) West Java.

#### Alternating target’s group

In this scenario, we change the target of age-groups in certain ways that is shown in Fig.21(a). High rate vaccination in the first month is given to active and older people, ages 20 and older. Fig.21(b) and Fig.22 describe the dynamics of total number *Q*(*t*) and *D*(*t*) of all age-groups obtained using this scenario in Jakarta, Banten, and West Java. Compared to those obtained by prioritizing active and older people, this scenario resulted the dynamics of active cases having thicker tail. This argument can be seen in every region we observed. Thus, this scenario is not likely preferable than the first scenario prioritizing the active and older people.

**Figure 21.**
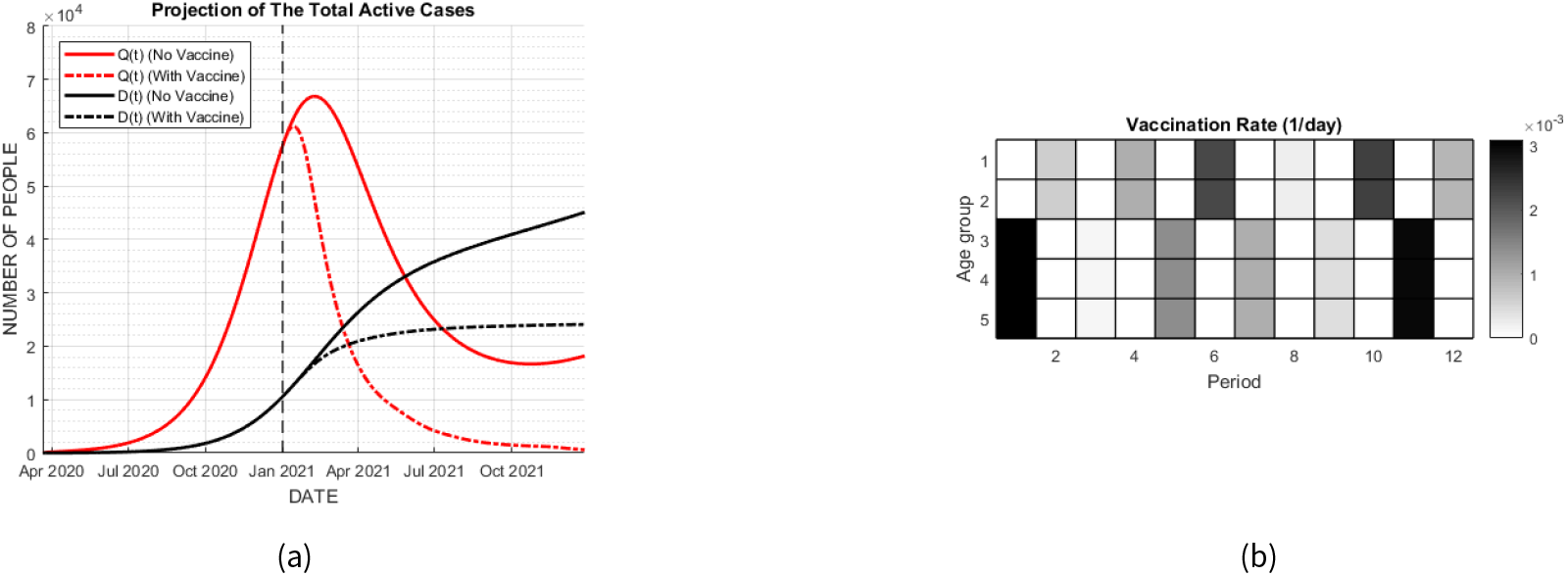
Alternating vaccination by switching vaccination target on each period **??** Vaccination schedule; (b) Dynamics of *Q*(*t*) and *D*(*t*) in Jakarta

**Figure 22.**
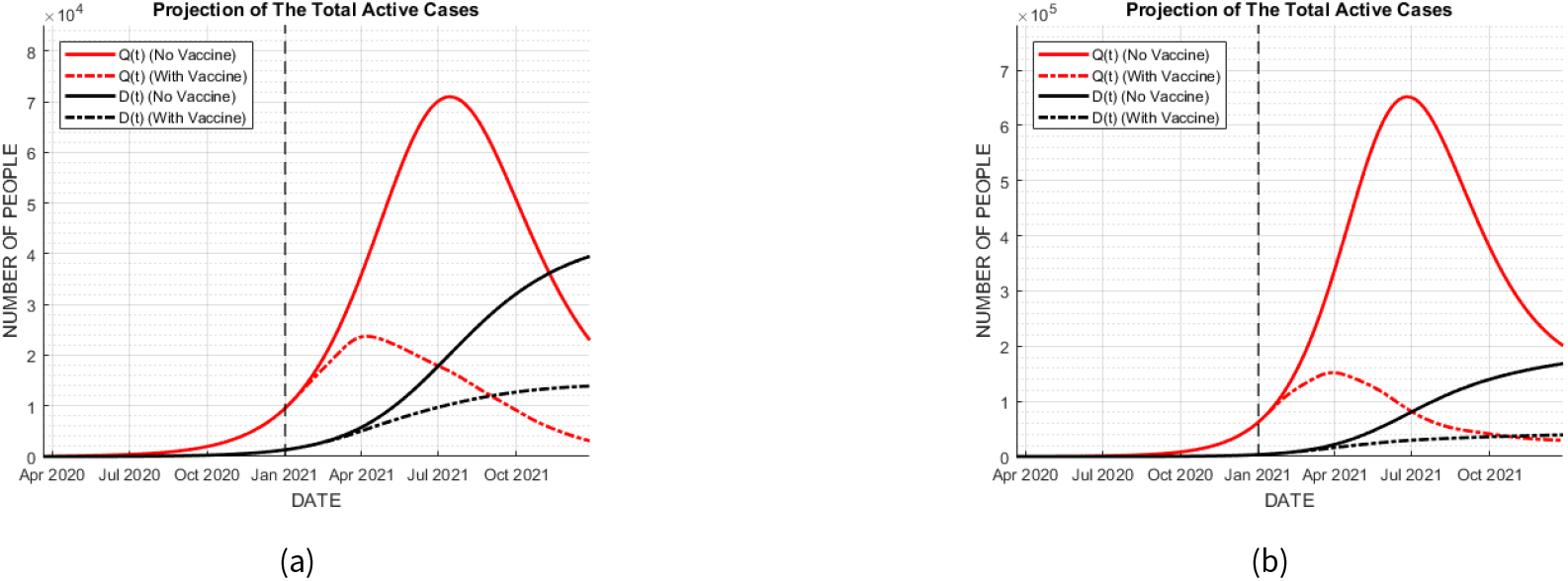
Alternating vaccination target on each period in (a) Banten; (b) West Java.

#### Only the active people

The Indonesia government is planning to conduct the vaccination only to the group of active people only, ages 20-49. We do the simulation in this scenario. Fig.23 depicts the dynamics of *Q*_*i*_ (*t*) once the third age-group is vaccinated. On the other hand, we also provide the dynamics of active cases that resulted from the other three scenarios as a comparison. Fig.24 shows that the vaccination prioritizing the active and older people is more preferable due to its significant reduction of cases. This argument is also valid to the results given in Banten and West Java as depicted by Fig.25.

**Figure 23.**
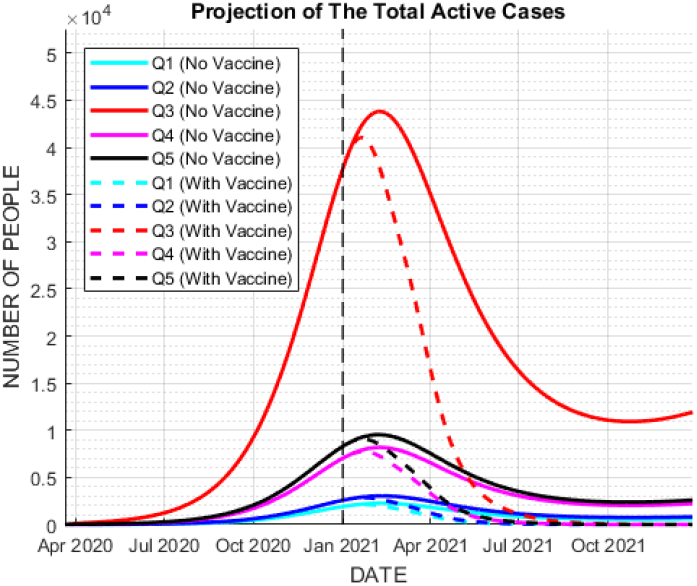
Dynamics of *Q*_*i*_ (*t*) once the third group of age in Jakarta is vaccinated

**Figure 24.**
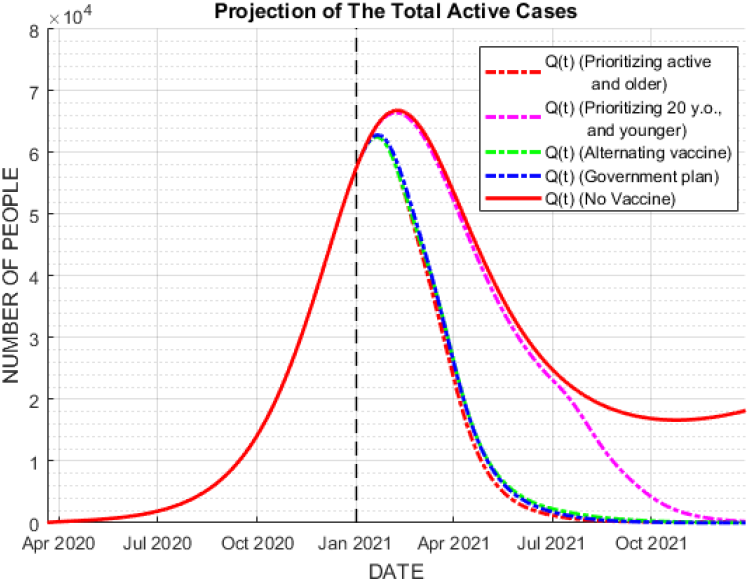
Comparison of *Q*(*t*) of all the observed vaccination scenarios in Jakarta

**Figure 25.**
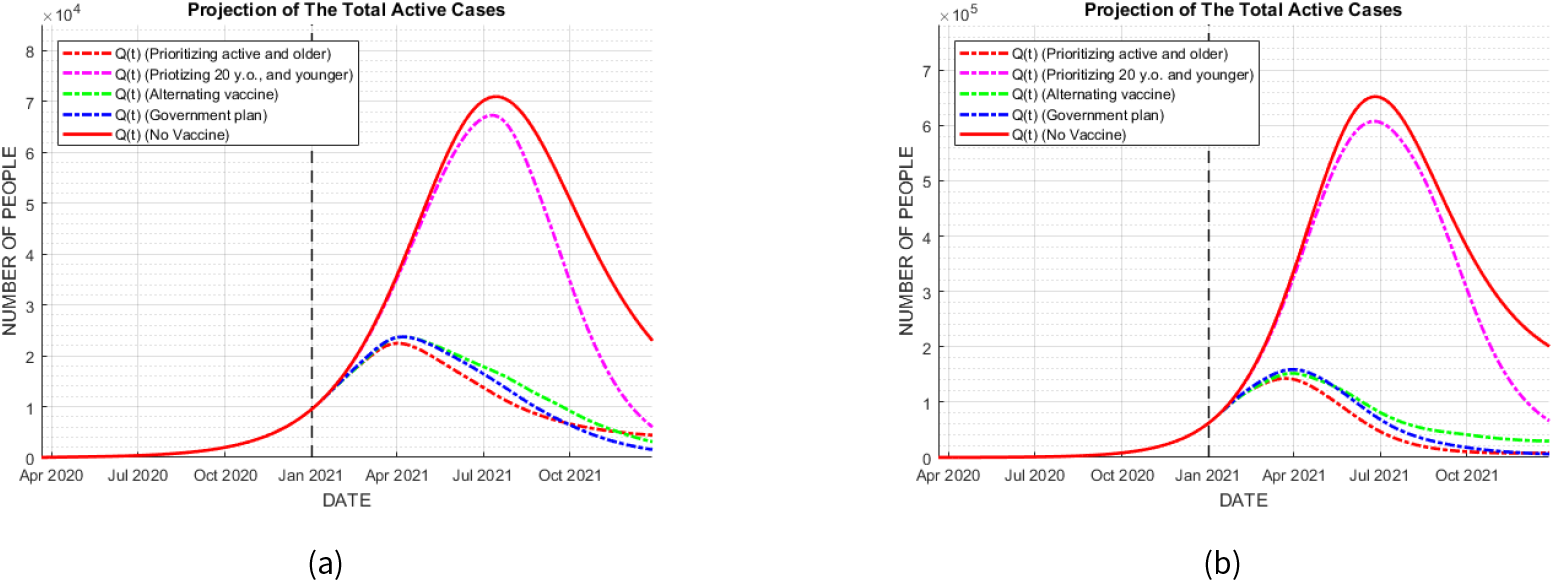
Comparison of *Q*(*t*) of all the observed vaccination scenarios in (a) Banten; (b) West Java.

**Figure 26.**
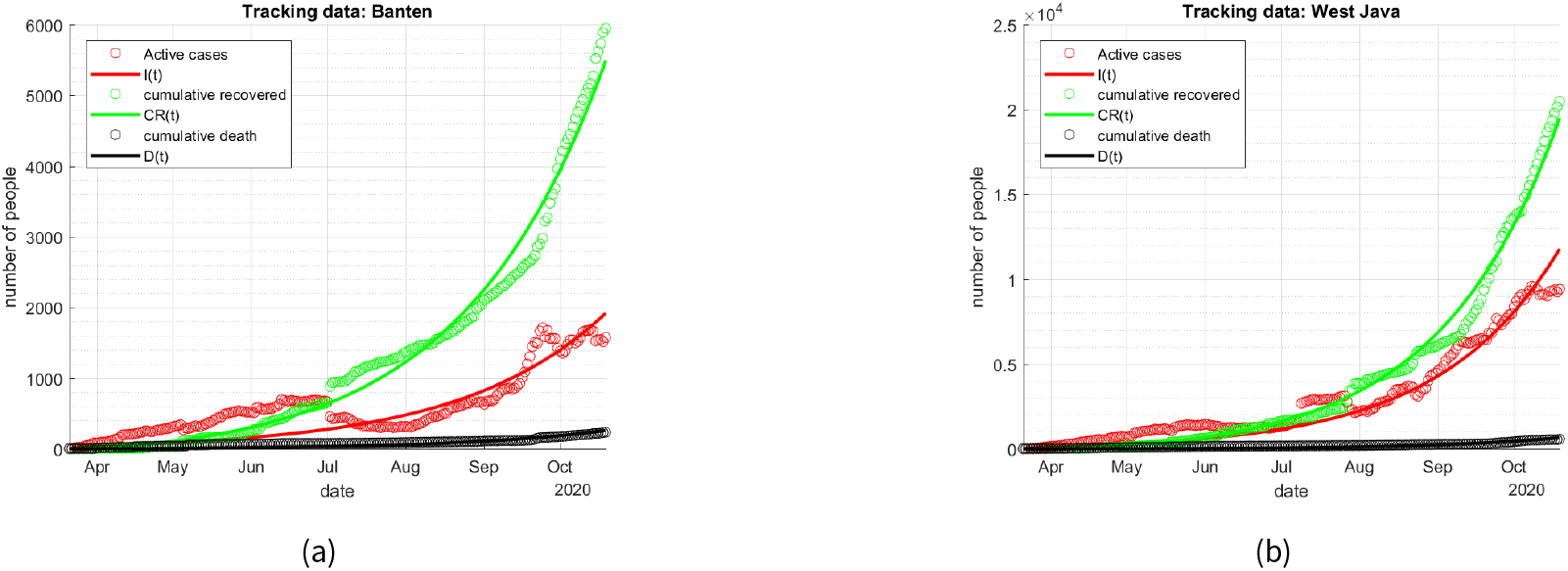
Fitting results of the non-age-structured model in: (a) Banten; (b) West Java

## Discussion

The simulations show that the vaccination scenario by first targeting active and older adults (above 50) (group 3-5) is better than other scenarios. The numbers of active cases and also total deaths were decreased significantly. In the result of scenario 2, the decrease was insignificant, because the transmission rate of virus among 20-year-old and younger people was much lower than of other age groups, as shown in Table 3.

After conducting the simulation by targeting the active people only, the dynamics from other age-groups on the number of active cases, *Q*_*i*_ (*t*), *i* ≠ 3, were also decreasing once people in the third age-group was vaccinated. It is logic because the key of the transmission is contact among people. The active people have larger access to people of other age-groups so the transmission across age groups is highly possible. Once the biggest source of infectious people is vaccinated, it affects the number of active cases on the other group of age.

## 7 Conclusions and Recommendations

Having constructed the SIQRD for non-age and age structures models and developed several scenarios on the implementation of vaccination program, we concluded our work by the following findings.

- The modification of the SIQRD model by adding the reinfection factor affected the behavior of the COVID-19 spread, where possibilities of more than one peaks of outbreak existed. The amplitude of active cases on the second or later peak of outbreaks was not as big as the first peak. The total number of deaths will increase over time when the vaccine is not available,
- The proper timing of vaccine implementation is in the early stage of the pandemic. This implementation will suppress the number of active cases immediately, and consequently the total deaths. After the number of active cases reaches its peak, the implementation of the vaccination program will not reduce the total deaths significantly.
- The vaccination should be implemented consistently following the planned schedule for a certain period. The vaccination’s implementation for only one or two months will not reduce the number of infectious persons, and eventually it will fail to prevent the second and more peaks of outbreak.
- The prioritization of the active and older adults (above 50) for vaccination over others and prioritization of active people only will significantly reduce the total deaths.

### A Graphs of non age-structured model

For provinces Banten and West Java, the comparisons of simulated and real data are following.

Those figures above are depicted by the estimation results of *β, γ, δ*, and *I*(*t* _0_) for Banten and West Java. The obtained parameters of the non age-structured model are given by the following table.

### B Estimated values of *β*_*i*_

We provide tables representing Obtained values *β*_*i*_ for the age-structured model of Banten and West Java.

**Table 8.** Parameter estimation of the non-age-structured model for Jakarta

| Region | $\beta$ | $\gamma$ | $\delta$ | $I(t_0)$ | $R_0$ |
| --- | --- | --- | --- | --- | --- |
| Banten | 0.5968 | 0.0520 | 0.0024 | 9 | 1.0447 |
| West Java | 0.6016 | 0.0354 | 0.0012 | 20 | 1.0528 |

**Table 9.** Obtained values *β*_*i*_ for the age-structured model of Banten and West Java

| Region | $\beta_1$ | $\beta_2$ | $\beta_3$ | $\beta_4$ | $\beta_5$ |
| --- | --- | --- | --- | --- | --- |
| Banten | 0.0253 | 0.0358 | 0.1131 | 0.1703 | 0.4525 |
| West Java | 0.0262 | 0.0371 | 0.1172 | 0.1765 | 0.4690 |

### C Vaccination scheme

We provide tables representing the estimation results of vaccination scheme for Banten and West Java with several starting times of vaccination.

1. Banten
  - Vaccination in October 2020;
  - Vaccination in January 2021;
  - After the peak of outbreak.
  - West Java
  - Vaccination in October 2020;
2. West Java
  - Vaccination in January 2021;
  - After the peak of outbreak.

**Table 10.**
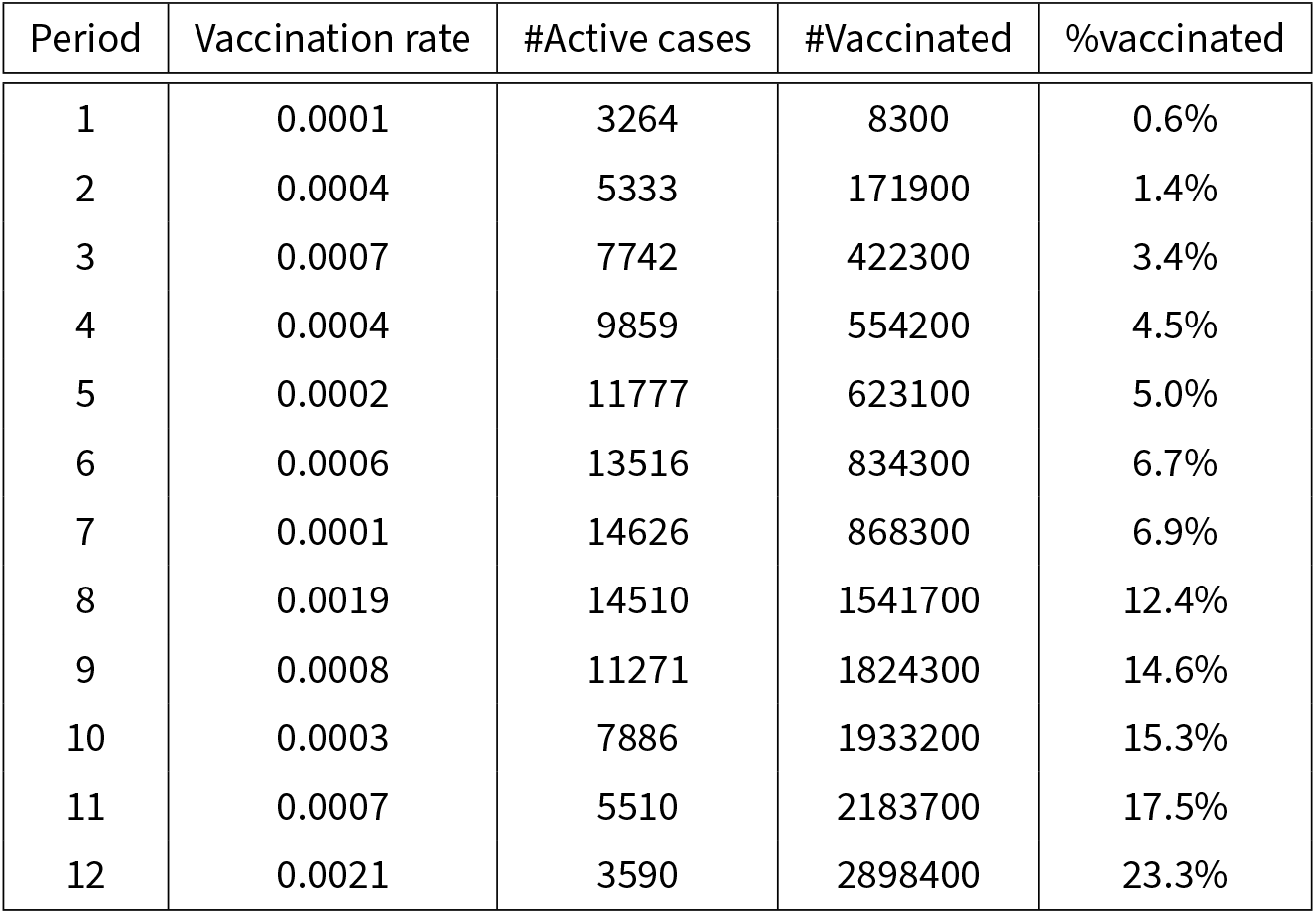
Estimation results of vaccination scheme starting Oct 2020 in Banten

**Table 11.** Estimation results of vaccination scheme starting Jan 2021 in Banten

| Period | Vaccination rate | #Active cases | #Vaccinated | %vaccinated |
| --- | --- | --- | --- | --- |
| 1 | 0.0007 | 11158 | 242500 | 2.1% |
| 2 | 0.0002 | 15906 | 318500 | 2.5% |
| 3 | 0.0012 | 19364 | 767000 | 6.2% |
| 4 | 0.0002 | 19984 | 842800 | 6.7% |
| 5 | 0.0001 | 19731 | 892100 | 7.2% |
| 6 | 0.0014 | 19468 | 1042300 | 8.4% |
| 7 | 0.0006 | 18434 | 1267200 | 10.1% |
| 8 | 0.0002 | 16524 | 1354600 | 10.8% |
| 9 | 0.0008 | 14436 | 1635000 | 13.1% |
| 10 | 0.0004 | 11873 | 1782800 | 14.3% |
| 11 | 0.0006 | 9512 | 1996200 | 16.1% |
| 12 | 0.0004 | 7474 | 2130200 | 17.1% |

**Table 12.** Estimation results of vaccination scheme starting after the peak of outbreak in Banten

| Period | Vaccination rate | #Active cases | #Vaccinated | %vaccinated |
| --- | --- | --- | --- | --- |
| 1 | 0.0001 | 70842 | 3000 | 0.1% |
| 2 | 0.0002 | 58624 | 68800 | 0.5% |
| 3 | 0.0003 | 47501 | 176900 | 1.4% |
| 4 | 0.0002 | 37728 | 231600 | 1.9% |
| 5 | 0.0001 | 30432 | 259600 | 2.1% |
| 6 | 0.0002 | 25370 | 353500 | 2.8% |
| 7 | 0.0003 | 21704 | 367500 | 2.9% |
| 8 | 0.0008 | 18818 | 655600 | 5.3% |
| 9 | 0.0004 | 15324 | 790200 | 6.3% |
| 10 | 0.0001 | 12330 | 838000 | 6.7% |
| 11 | 0.0003 | 10262 | 955200 | 7.7% |
| 12 | 0.0008 | 8503 | 1258900 | 10.1% |

**Table 13.**
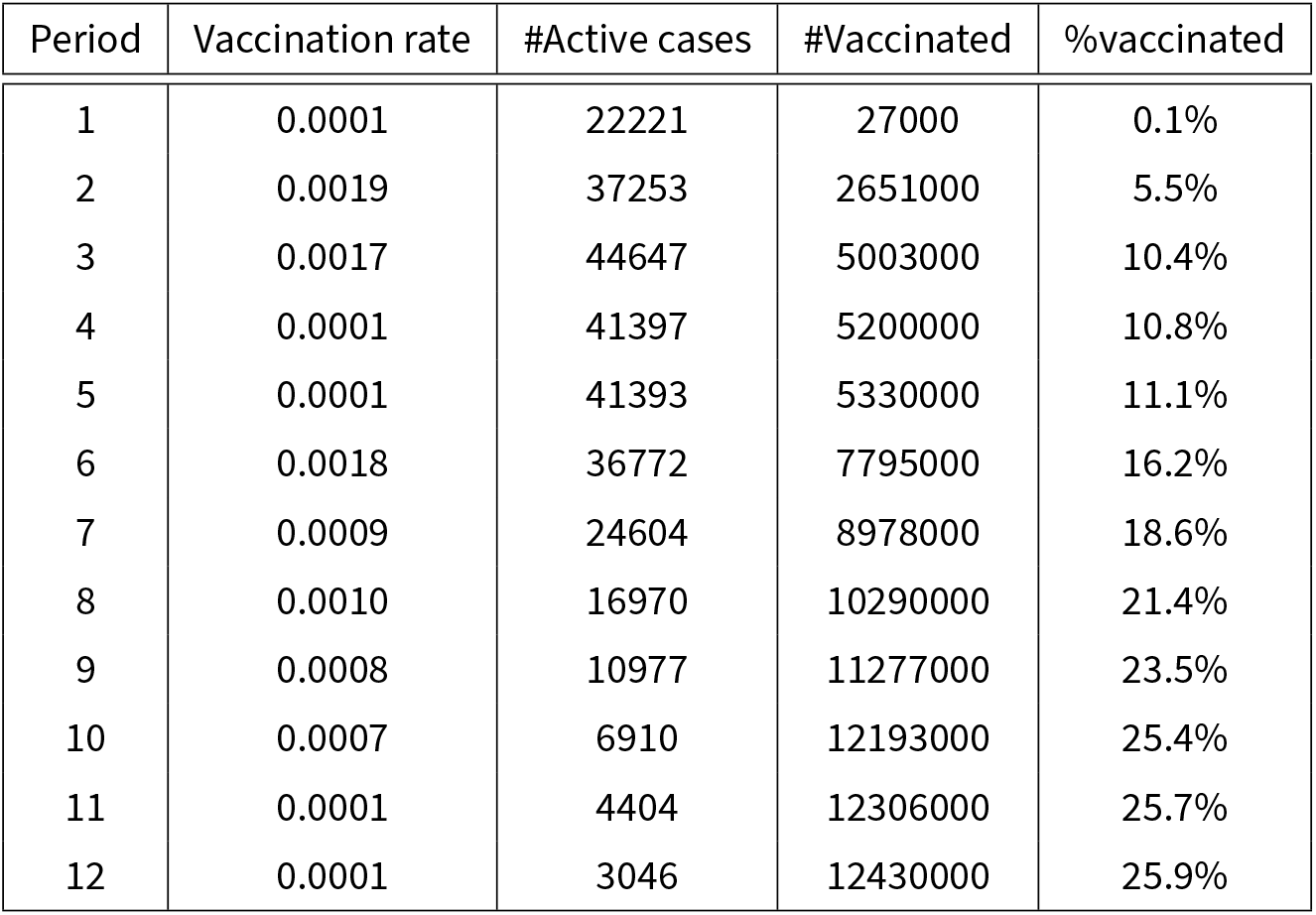
Estimation results of vaccination scheme starting on Oct 2020 in West Java

**Table 14.** Estimation results of vaccination scheme starting on Jan 2021 in West Java

| Period | Vaccination rate | #Active cases | #Vaccinated | %vaccinated |
| --- | --- | --- | --- | --- |
| 1 | 0.0010 | 92140 | 2187000 | 4.5% |
| 2 | 0.0008 | 117610 | 3333000 | 6.9% |
| 3 | 0.0012 | 122050 | 4921000 | 10.2% |
| 4 | 0.0006 | 107180 | 5678000 | 11.8% |
| 5 | 0.0001 | 88060 | 5879000 | 12.2% |
| 6 | 0.0011 | 71690 | 7354000 | 15.3% |
| 7 | 0.0010 | 54550 | 8623000 | 17.9% |
| 8 | 0.0005 | 38940 | 9295000 | 19.3% |
| 9 | 0.0002 | 27710 | 9533000 | 19.8% |
| 10 | 0.0008 | 20560 | 10584000 | 22.0% |
| 11 | 0.0002 | 15610 | 10804000 | 22.5% |
| 12 | 0.0001 | 12710 | 10829000 | 22.7% |

**Table 15.** Estimation results of vaccination scheme starting after the peak of outbreak in West Java

| Period | Vaccination rate | #Active cases | #Vaccinated | %vaccinated |
| --- | --- | --- | --- | --- |
| 1 | 0.0001 | 676430 | 22900 | 0.1% |
| 2 | 0.0004 | 548920 | 495200 | 1.1% |
| 3 | 0.0006 | 409940 | 1235900 | 2.5% |
| 4 | 0.0003 | 286400 | 1621700 | 3.3% |
| 5 | 0.0002 | 197270 | 1822400 | 3.8% |
| 6 | 0.0005 | 139270 | 2461500 | 5.1% |
| 7 | 0.0001 | 101840 | 2561500 | 5.3% |
| 8 | 0.0014 | 76470 | 4480700 | 9.3% |
| 9 | 0.0007 | 53650 | 5363600 | 11.1% |
| 10 | 0.0003 | 25220 | 5697800 | 11.8% |
| 11 | 0.0006 | 17400 | 6482300 | 13.4% |
| 12 | 0.0016 | 5023 | 8558300 | 17.8% |

## Data Availability

Data are provided in an online respiratory.

https://github.com/agusisma/covidvaccine

